# A Yeast-Based Platform for Oral Delivery of a Dual GLP-1/GIP Receptor Agonist: Engineering and Preliminary Clinical Evaluation

**DOI:** 10.64898/2026.10.22.26365662

**Authors:** Kwang-Chul Kwon, Olga Gaidarenko, Nathan Chang, Cory Henderson

## Abstract

Oral delivery of peptides and other biologics are limited by enzymatic degradation and poor intestinal permeability. We developed a *Saccharomyces cerevisiae*–based platform expressing pro-DALAP, a fusion precursor of a dual GLP-1/GIP receptor agonist. The construct incorporates maltose-binding protein, a GM1-binding peptide, a cell-penetrating peptide, furin cleavage sites, and an albumin-binding domain, designed to support peptide stability, epithelial uptake, proteolytic activation, and prolonged systemic exposure. CRISPR/Cas12a-mediated integration of three expression cassettes into a protease-deficient yeast strain yielded 4.05 ± 0.31 mg pro-DALAP/g dry biomass, compared with 1.27 ± 0.21 mg/g in the single-copy strain, without detectable changes in cellular morphology or growth under the conditions evaluated. In vitro assays confirmed GM1 binding, furin-mediated processing, and activation of both GLP-1 and GIP receptors by DALAP. Preliminary intravenous toxicology studies in rats identified no treatment-related adverse findings at doses up to 300 µg/kg/day for seven days. A 12-week, open-label, randomized controlled trial evaluated enteric-coated capsules containing lyophilized yeast expressing pro-DALAP against dietary guidance alone. The baseline cohort comprised 98 generally healthy adults who were overweight. An exploratory interim week-8 analysis involving eight participants per group showed numerically greater reductions in body weight (−2.29% vs. +0.14%) and waist circumference (−6.34% vs. −2.32%) in the intervention group, although neither comparison was statistically significant. These larger numerical differences were not reproduced in the completed analysis. Over the 12-week study, the intervention group showed modest numerical reductions in body weight and waist circumference. At week 12, mean percentage changes from baseline in body weight were −0.76% in the intervention group and −0.27% in the control group; corresponding changes in waist circumference were −3.60% and −3.69%. These descriptive trends did not translate into statistically significant between-group differences for either outcome. HbA1c reduction favored the control group. Among 40 intervention-group survey respondents, 37 reported no side effects. These findings support yeast-based production and in vitro functionality of pro-DALAP and provide preliminary observations on safety and efficacy. Validated pharmacokinetic measurements and adequately powered, blinded, placebo-controlled trials with longer follow-up periods are needed to determine systemic exposure, and evaluate sustained benefits, potentially supporting future clinical applications.

## Introduction

Peptide-based therapeutics, particularly incretin mimetics targeting GLP-1 and GIP receptors, have demonstrated significant clinical efficacy in obesity and metabolic disorders [1, 2]. However, their widespread use remains limited by the requirement for subcutaneous administration and the challenges associated with oral delivery, including enzymatic degradation and poor intestinal absorption [3].

Over the past two decades, incretin-based therapies have evolved significantly, beginning with the approval of exenatide in 2005 as the first GLP-1 receptor agonist requiring twice-daily injection for patients with type 2 diabetes [4]. Subsequent advances focused on improving pharmacokinetics and patient compliance, leading to the development of long-acting formulations such as liraglutide and lixisenatide (once-daily injection) [5, 6] and once-weekly agents, including albiglutide [7], dulaglutide [8] and semaglutide [9]. A major milestone was achieved with the approval of oral semaglutide in 2019, which utilizes a chemical absorption enhancer to facilitate gastric uptake [10, 11]. More recently, dual and triple agonists, such as tirzepatide and retatrutide, have demonstrated substantial improvements in weight loss efficacy by targeting multiple metabolic pathways [2, 12].

Despite these advances, oral delivery of peptide therapeutics remains inefficient and formulation-dependent. Current strategies, including chemical modification, permeation enhancers, and nanoparticle-based delivery systems, have shown partial success but are often limited by low and variable bioavailability [3]. As a result, achieving efficient oral bioavailability while maintaining biological activity remains a critical unmet need.

Among current oral peptide approaches, oral semaglutide (Rybelsus) represents a notable advance, relying on co-formulation of the synthetic peptide with the absorption enhancer sodium N-(8-[2-hydroxybenzoyl] amino) caprylate (SNAC) to facilitate transcellular uptake in the stomach [10]. However, this strategy is associated with several limitations, including low and variable bioavailability, strict dosing requirements, and dependence on synthetic formulation components [11, 13]. These limitations highlight the need for alternative strategies that can achieve effective oral delivery while maintaining biological activity and scalability.

In addition, many current incretin therapeutics, including semaglutide and tirzepatide, incorporate non-natural amino acid substitutions and fatty acid conjugation to enhance proteolytic stability and extend plasma half-life [9, 14]. While these modifications effectively improve pharmacokinetic properties, they may also introduce risks related to altered immunogenicity, off-target interactions, and prolonged systemic exposure. Such structural modifications may contribute to the gastrointestinal adverse effects commonly associated with incretin-based therapies (e.g., nausea, vomiting, diarrhea, constipation, and bloating) [1, 2] and may complicate dose optimization owing to prolonged systemic exposure.

In contrast, the platform in this study employs a biologically engineered delivery mechanism that integrates peptide stabilization, epithelial targeting, and intracellular activation within a single fusion construct. Rather than relying on chemical permeation enhancers, transcytosis is facilitated through ligand-receptor interactions, specifically, binding of the GM1-targeting peptide to GM1 as well as electrostatic interactions between the positively charged cell-penetrating peptide and the negatively charged extracellular matrix of epithelial cells, followed by proteolytic activation via endogenous furin. This approach enables a more physiologically driven transport process and eliminates the need for synthetic absorption enhancers and non-natural amino acid modifications.

Furthermore, while oral semaglutide requires complex chemical manufacturing and formulation [9, 10], our system leverages yeast-based production, offering a scalable and potentially lower-cost alternative. Notably, the current implementation as a dietary supplement enables real-world validation and rapid deployment while maintaining a pathway toward therapeutic development. Accordingly, our platform avoids synthetic chemical modifications and instead relies on biologically mediated stabilization and activation, reducing risks associated with prolonged exposure and non-natural structural elements.

In this study, we establish yeast as a dual platform for protein expression and oral delivery. *Saccharomyces cerevisiae* is a well-characterized unicellular eukaryotic microorganism with a fully sequenced genome and a long history of safe use in food and biotechnology [15, 16]. It has been extensively utilized in the production of fermented foods and beverages, including bread, beer, wine, and dairy products, and is therefore routinely consumed by humans in substantial quantities [16, 17]. This long-standing dietary exposure, combined with decades of scientific investigation, has established *S. cerevisiae* as one of the most widely accepted and trusted organisms for industrial and nutritional applications [16]. In addition, it serves as a key model organism in molecular and cellular biology, with a deeply characterized genetic system that enables precise and predictable genetic modification [15, 18]. The safety of *S. cerevisiae* is supported by multiple global regulatory authorities. The U.S. Food and Drug Administration (FDA) has approved dried yeast and numerous yeast-derived components, including proteins, glycans, and extracts, for use in food, and has designated *S. cerevisiae* as Generally Recognized as Safe (GRAS) in multiple notices [19, 20]. Similarly, the Joint FAO/WHO Expert Committee on Food Additives (JECFA), in specifications originating from its 1971 evaluation of carbohydrases, describes Saccharomyces species as traditionally used in food manufacture [21], while the U.S. Environmental Protection Agency (EPA) includes it among organisms eligible for regulatory exemptions due to its lack of pathogenicity and toxicity [16]. In Europe, the European Food Safety Authority (EFSA) has assigned *S. cerevisiae* Qualified Presumption of Safety (QPS) status [22], further supporting its safe use in food and feed without the need for extensive additional safety evaluation. Collectively, these regulatory designations reflect a strong consensus regarding the safety profile of this organism.

We developed a biologically engineered oral delivery system using *S. cerevisiae*. The functional peptide (DALAP) was expressed as a fusion protein (pro-DALAP) incorporating several functional modules: (i) a GM1-binding peptide and a cell-penetrating peptide to facilitate intestinal epithelial uptake, (ii) maltose-binding protein (MBP) to enhance solubility and protect DALAP from proteolysis, (iii) furin cleavage sites to release active DALAP into circulation during transcytosis, and (iv) an albumin-binding domain to extend systemic half-life of DALAP.

We hypothesized that this multifunctional fusion strategy would support oral delivery, proteolytic activation, and systemic exposure of the active peptide, DALAP. To investigate the platform’s potential for oral use in humans, we conducted a 12-week, open-label, randomized controlled trial evaluating enteric-coated capsules containing lyophilized yeast expressing pro-DALAP in generally healthy adults who were overweight. The study assessed percentage changes from baseline in body weight and waist circumference, changes in glycemic control, and participant-reported outcomes and tolerability. Both body weight and waist circumference showed overall downward trends during the 12-week study. At week 12, the intervention group showed a modestly greater numerical reduction in body weight than the control group, whereas reductions in waist circumference were similar between groups. Neither between-group difference was statistically significant. Here, we present the design, genome engineering, and functional characterization of the yeast-based platform alongside the completed clinical findings, providing a basis for further pharmacokinetic investigation and longer, adequately powered, placebo-controlled studies.

## Materials and Methods

### Generation of a Single-Copy Yeast Transformant

Single-copy integration was performed using a homology-directed repair (HDR) strategy at the *URA3* locus. Yeast transformation was performed using the lithium acetate/single-stranded carrier DNA/polyethylene glycol (LiAc/SS carrier DNA/PEG) method described by Gietz and Schiestl (2007) [23], with modifications. Briefly, cells were grown overnight in YPD medium at 30 °C with shaking and diluted in fresh YPD to an OD₆₀₀ of approximately 0.1. Cultures were grown to mid-log phase (OD₆₀₀ ≈ 0.4–0.8), harvested by centrifugation, washed with sterile water, and resuspended in 100 mM LiAc. Cells were then combined with a transformation mixture containing 50% (w/v) PEG 3350, 1 M LiAc, heat-denatured salmon sperm DNA as a carrier, and the appropriate DNA fragments. The mixtures were mixed thoroughly and heat-shocked at 42 °C for 45 min, followed by recovery and plating on selective agar media.

First, ura3 knockout derivatives of *Saccharomyces cerevisiae* strains were generated by transformation with a 100-bp DNA fragment prepared by annealing and extension of primers 102 and 103. Following overnight recovery in YPD medium, transformed cells were plated on synthetic defined (SD) medium supplemented with uracil and 5-fluoroorotic acid (5-FOA). Colonies were screened by colony PCR to distinguish the intended *URA3* disruption from spontaneous 5-FOA-resistant mutants.

The *ura3* knockout strains were subsequently cotransformed with three DNA fragments designed for in vivo assembly of the pro-DALAP expression construct and a functional *URA3* selection marker, followed by HDR-mediated integration at the disrupted *URA3* locus. Transformants were selected on SD medium lacking uracil and initially screened by colony PCR using primers 163 and 164 to amplify the integrated pro-DALAP sequence for sequencing. Integration was confirmed by sequencing, and protein expression was verified by Western blotting. The final strain was cryopreserved as a glycerol stock. Primer sequences are listed in **Supplementary Table 1.**

### CRISPR/Cas12a-Mediated Genome Engineering

Genome engineering in *Saccharomyces cerevisiae* was performed using a CRISPR/Cas12a-assisted homologous recombination strategy. Guide RNAs (gRNAs) targeting specific genomic loci, including *URA3* and additional integration sites (II-1, XI-3, and YPRCτ3), were expressed from plasmids carrying a *URA3* selection marker. The II-1, XI-3, and YPRCτ3 integration sites were selected based on the strategy described by Bennis et al. (2023) [18], who used these nonessential chromosomal regions for targeted integration and multiplex genome editing. Donor DNA fragments containing the expression cassette were generated by PCR with locus-specific homology arms to enable targeted integration. Primer sequences used for donor DNA amplification, construct assembly, and verification of genomic modifications are listed in **Supplementary Table 1.**

Yeast transformation was performed using the LiAc/PEG-mediated method described above. Donor DNA fragments (linear PCR products) and CRISPR/Cas12a gRNA plasmids (100–300 ng each) were cotransformed into yeast cells. Transformants were selected on appropriate media and screened by PCR. To remove the editing plasmids following genome editing, strains were passaged in nonselective medium and subjected to counterselection on 5-FOA plates to select for loss of the *URA3*-marked plasmids. Successful genomic integrations and gene disruptions were confirmed by PCR and/or sequencing. For construction of multicopy strains, multiplexed CRISPR/Cas12a editing was performed by cotransforming multiple donor DNA fragments with the corresponding gRNAs targeting distinct genomic loci.

To minimize potential degradation of pro-DALAP during expression in yeast, a protease-deficient *Saccharomyces cerevisiae* host was generated by disrupting *PEP4* and *PRB1* using CRISPR/Cas12a. The resulting strain was used for both HDR-mediated single-copy integration and CRISPR/Cas12a-mediated multiplex integration.

### Protein Extraction, Western Blotting, and Quantification of Recombinant Protein Expression

Total protein was extracted from engineered *Saccharomyces cerevisiae* strains using a lysis buffer consisting of 1× PBS supplemented with protease inhibitor cocktail, 5 mM EDTA, and 1,10-phenanthroline. For dried yeast samples, cells were first mechanically disrupted using a mortar and pestle prior to extraction. Samples were resuspended in extraction buffer at a defined ratio (typically 10 mg biomass per 300–600 µL buffer, depending on sample type) and lysed by sonication (5 s on/5 s off cycles, repeated four times) on ice. Clarified lysates were used for downstream analysis. Total protein concentration was determined using a Bradford assay with bovine serum albumin (BSA) as a standard. For immunoblot analysis, protein samples were normalized to equal concentrations, mixed with SDS sample buffer, and heat-denatured at 80 °C for 3 min. Proteins were separated by Tris–SDS polyacrylamide gel electrophoresis (typically 10% gels) and transferred onto nitrocellulose or PVDF membranes using a wet transfer system (80 V, ∼50 min). Membranes were blocked with 3–5% (w/v) non-fat dry milk in PBST (PBS containing 0.1% Tween-20) and incubated with an anti-MBP monoclonal primary antibody (New England Biolabs, catalog no. E8032S; approximately 1:10,000 dilution), followed by an HRP-conjugated goat anti-mouse IgG (H+L) secondary antibody (Thermo Fisher Scientific, catalog no. 31430; approximately 1:20,000 dilution). Chemiluminescent signals were developed and imaged using an iBright 1500 imaging system (Thermo Fisher Scientific). For quantitative analysis, purified MBP standard protein was loaded alongside samples at defined amounts (typically 4–32 ng per lane) to generate a calibration curve within the linear detection range. Band intensities were quantified using iBright analysis software, and background-corrected signal volumes were used to construct a standard curve. The amount of recombinant protein in each sample was determined by interpolation from the MBP standard curve. Expression levels were calculated by normalizing the quantified protein amount to the total extraction volume and input biomass, and were reported as micrograms of recombinant protein per gram of dry cell weight (µg/g DW) [24]. Specifically, expression levels were derived using the relationship:

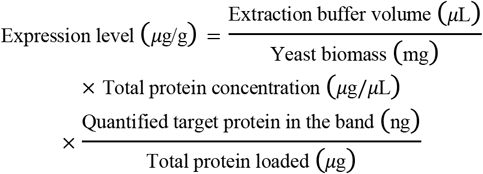

All measurements were performed using independent biological replicates, and values are reported as mean ± standard error where applicable.

### GM1-Ganglioside Binding ELISA Assay

GM1 binding of pro-DALAP was evaluated by ELISA adapted from Kwon et al. (2018) [24], with modifications for yeast-derived protein extracts and pro-DALAP detection. Ninety-six–well microtiter plates were coated with GM1-ganglioside (Sigma, G-7641) at a concentration of 3 µg/mL in bicarbonate buffer (15 mM Na₂CO₃, 35 mM NaHCO₃, pH 9.6), with 100 µL added per well and incubated overnight at 4°C. Following coating, plates were washed three times with PBST (PBS containing 0.1% Tween-20) and blocked with 200 µL of 3% (w/v) non-fat dry milk in PBST for 1–2 h at 37°C. After blocking, plates were washed again three times with PBST. Total soluble protein extracts from transformed and untransformed yeast were prepared in extraction buffer (1× PBS supplemented with protease inhibitor cocktail and 5 mM EDTA). Samples were normalized to a final concentration of 1 µg/µL and serially diluted (1/2, 1/4, and 1/8). A volume of 100 µL per well was added and incubated either overnight at 4°C or for 2 h at 37°C. As negative controls, 1% bovine serum albumin (BSA) and 1% skim milk in PBS were included. After incubation, plates were washed three times with PBST and then incubated with anti-MBP primary antibody diluted 1:10,000 in PBST containing milk for 1 h at 37°C. Plates were washed again and incubated with horseradish peroxidase (HRP)-conjugated anti-mouse secondary antibody (1:5,000 dilution) for 1 h at 37°C. Following a final wash step, 100 µL of TMB substrate was added to each well, and color development was allowed to proceed in the dark for 10–20 min at 37°C. The reaction was stopped by adding 50 µL of 2 N H₂SO₄, and absorbance was measured at 450 nm using a microplate reader.

### GPCR Activation Assay and Assessment of Albumin Binding

Receptor activation was evaluated using a G protein–coupled receptor (GPCR) signaling assay coupled to intracellular cyclic adenosine monophosphate (cAMP) production. Human embryonic kidney (HEK293) cells stably expressing either the GLP-1 receptor or GIP receptor were used for all experiments. Cells were cultured under standard conditions and seeded into multiwell plates prior to assay. Intervention compounds, including active EV1-Peptide, comparator agonists (i.e., Exendin-4 and GIP), were prepared in assay buffer and applied to cells over a range of concentrations to generate dose–response curves. To assess albumin binding effects, compounds were incubated in the presence or absence of human serum albumin (HSA) prior to addition to cells. Following ligand stimulation, intracellular cAMP levels were quantified using a luminescence-based detection system linked to adenylate cyclase activity. Luminescence signals were measured using a microplate reader and normalized to control conditions. Concentration–response curves were fitted using nonlinear regression, and half-maximal effective concentration (EC₅₀) values were calculated. Data are presented as log₁₀(EC₅₀) values. All experiments were performed with independent biological replicates, and values are reported as mean ± standard error where applicable.

### Furin Cleavage Assay

Pro-DALAP was purified using amylose magnetic beads (New England Biolabs, catalog no. E8035S). Purified pro-DALAP was incubated with recombinant furin (New England Biolabs, catalog no. P8077S) in reaction buffer containing 50 mM Tris-HCl (pH 7.4), 150 mM NaCl, 4 mM CaCl₂, and 10 mM KCl, using buffer conditions similar to those described by Oleaga et al. (2021) [25]. Reactions were terminated by adding SDS sample buffer followed by heat denaturation. Reaction products were separated by SDS–PAGE and analyzed by Western blotting using an anti-MBP antibody, following the procedure described above.

### Growth Rate Determination

Growth kinetics of the parent, single-copy, and three-copy strains were evaluated under non-selective conditions. Cryopreserved glycerol stocks were first recovered by inoculation into YPD medium and passaged daily for three consecutive days (5 µL into 5 mL YPD) to minimize potential artifacts associated with recovery from cryopreservation. Cultures were then grown in YPD at 30 °C with shaking at 250 rpm. Following recovery, cells were diluted to an initial OD₆₀₀ of approximately 0.05 and incubated under the same conditions. Optical density at 600 nm (OD₆₀₀) was measured hourly over an 8-hour period using a spectrophotometer. All measurements were performed in biological triplicates. Maximum specific growth rates (µmax) were determined by plotting the natural logarithm of OD₆₀₀ values over time and applying linear regression to the exponential growth phase, using at least five consecutive data points. Data are presented as mean ± standard error (s.e.m.) from independent biological replicates.

### Intravenous Tolerability and Repeat-Dose Toxicity Study in Rats

An exploratory, non-GLP study was conducted at ITR Laboratories Canada Inc. (Baie-d’Urfé, Quebec, Canada; study No. 701915) to evaluate single-dose tolerability and short-term repeat-dose toxicity of DALAP, designated DA-LA-P in the study records. The study plan and amendments were accepted by ITR’s Animal Care Committee. Animal care followed Canadian Council on Animal Care and NIH guidelines, and procedures complied with ITR standard operating procedures.

Twelve male Sprague–Dawley rats obtained from Charles River Laboratories were acclimated for at least two weeks and allocated by body-weight-based block randomization to two phases of six animals each. Animals were approximately nine weeks old at treatment initiation and maintained under controlled environmental conditions with a 12-hour light/dark cycle and food and water available ad libitum, except during designated procedures.

Lyophilized DALAP (reported purity ≥95%) was dissolved in sterile water for injection and diluted with Dulbecco’s phosphate-buffered saline (DPBS). Formulations were filtered through a 0.22-µm membrane and used fresh or stored frozen. Vehicle consisted of sterile water and 1× DPBS (3:1, v/v). Formulation concentrations, homogeneity, and stability were not analytically verified.

In Phase I, groups of two rats received a single intravenous dose of 10, 100, or 300 µg/kg, administered sequentially by dose group to assess tolerability. Animals were observed for five days and necropsied on day 6. In Phase II, groups of two rats received vehicle or DALAP at 100 or 300 µg/kg/day for seven consecutive days, with terminal examinations on day 8. All injections were administered through the tail vein over 30–60 seconds at 10 mL/kg.

Assessments included mortality, clinical signs, body weight, food consumption, and terminal gross necropsy. Phase II additionally included hematology, clinical chemistry, organ weights, and histopathological examination of protocol-specified tissues and gross lesions. At termination, animals were fasted overnight, anesthetized with isoflurane, and euthanized by exsanguination.

Data were recorded using Provantis software and reported individually, with Phase II numerical results also summarized as means and standard deviations. No inferential statistical analyses were performed because of the small group sizes.

### Randomized Controlled Clinical Study

A 12-week, open-label, randomized, controlled, two-arm clinical study was conducted by Citruslabs to evaluate the effects of an oral formulation containing engineered *Saccharomyces cerevisiae* expressing pro-DALAP on body weight, glycemic control, and appetite-related outcomes. The study protocol and accompanying documents were approved by Argus Independent Review Board (AIRB; Tucson, Arizona) on December 6, 2024 (protocol No. 20692). The protocol specified compliance with International Council for Harmonisation Good Clinical Practice (ICH-GCP) guidelines and applicable U.S. regulations. All participants provided written informed consent before study-specific procedures.

The original enrollment target was 120 adults, with 60 participants per group. A documented amendment dated February 3, 2026, revised the target to 98 participants. The final baseline cohort comprised 98 participants, including 51 in the intervention group and 47 in the control group. The original protocol specified eligibility criteria of age ≥18 years, BMI 25–29.9 kg/m², concerns regarding weight management, appetite or food cravings, and blood sugar regulation, and generally good health without uncontrolled chronic disease. Key exclusion criteria included type 1 diabetes, specified metabolic or endocrine disorders, pharmacologically treated type 2 diabetes, recent use of medications or supplements affecting weight or glycemic control, pregnancy or breastfeeding, and conditions that could interfere with study participation.

Participants were assigned to the intervention or control group using a computerized randomization tool. The intervention group received oral capsules containing dried, powdered yeast expressing pro-DALAP, providing approximately 4.74 mg of pro-DALAP per day. The control group received no study product or placebo and followed standardized healthy eating recommendations based on American Diabetes Association guidance.

The study employed a hybrid design, combining remote questionnaires and home measurements with laboratory blood collection. Body weight and waist circumference were assessed at baseline and weeks 4, 6, 8, and 12 using digital scales and measuring tapes, respectively. Blood samples were collected for measurement of hemoglobin A1c (HbA1c) and estimated average glucose (eAG). The capsule dosing schedule, laboratory provider, blood collection time points, and detailed measurement procedures should be specified according to the final amended protocol and study records.

The protocol specified changes in body weight and HbA1c as primary outcomes. Secondary outcomes included participant-reported perceptions of appetite suppression, food cravings, weight management, blood sugar regulation, and overall well-being, assessed using structured questionnaires. Waist circumference was also assessed in the amended study.

The protocol specified an intention-to-treat analysis, including between-group comparisons and, where appropriate, within-group comparisons against baseline. Descriptive results were summarized using the available measurements at each visit, with the corresponding participant numbers reported. The final analysis population, handling of missing data, statistical tests, covariate adjustments, and measures of variability should be specified according to the completed statistical analysis report.

## Results

### Development of pro-DALAP

The two main incretin hormones, glucagon-like peptide-1 (GLP-1) and glucose-dependent insulinotropic polypeptide (GIP), enhance glucose-dependent insulin secretion from pancreatic β-cells [26]. GLP-1 also slows gastric emptying [26] and reduces food intake through gut–brain signaling pathways [27]. Contrary to the earlier concept of GIP promoting fat deposition, GIP receptor activation has been found to reprogram adipose tissue towards healthier energy expenditure and reduced fat accumulation [28, 29].

Endogenous glucose-dependent insulinotropic polypeptide (GIP) and glucagon-like peptide-1 (GLP-1) undergo rapid enzymatic inactivation, with circulating half-lives of approximately 5 and 2 minutes, respectively, limiting their duration of action in circulation [26, 30]. To combine their receptor activities within a single peptide and address susceptibility to proteolytic degradation, we designed a GIP-based dual agonist (DA) incorporating an Ala2-to-Gly substitution and a modified C-terminal region intended to support Trp-cage-like folding (**Figure 1A**).

**Figure 1.**
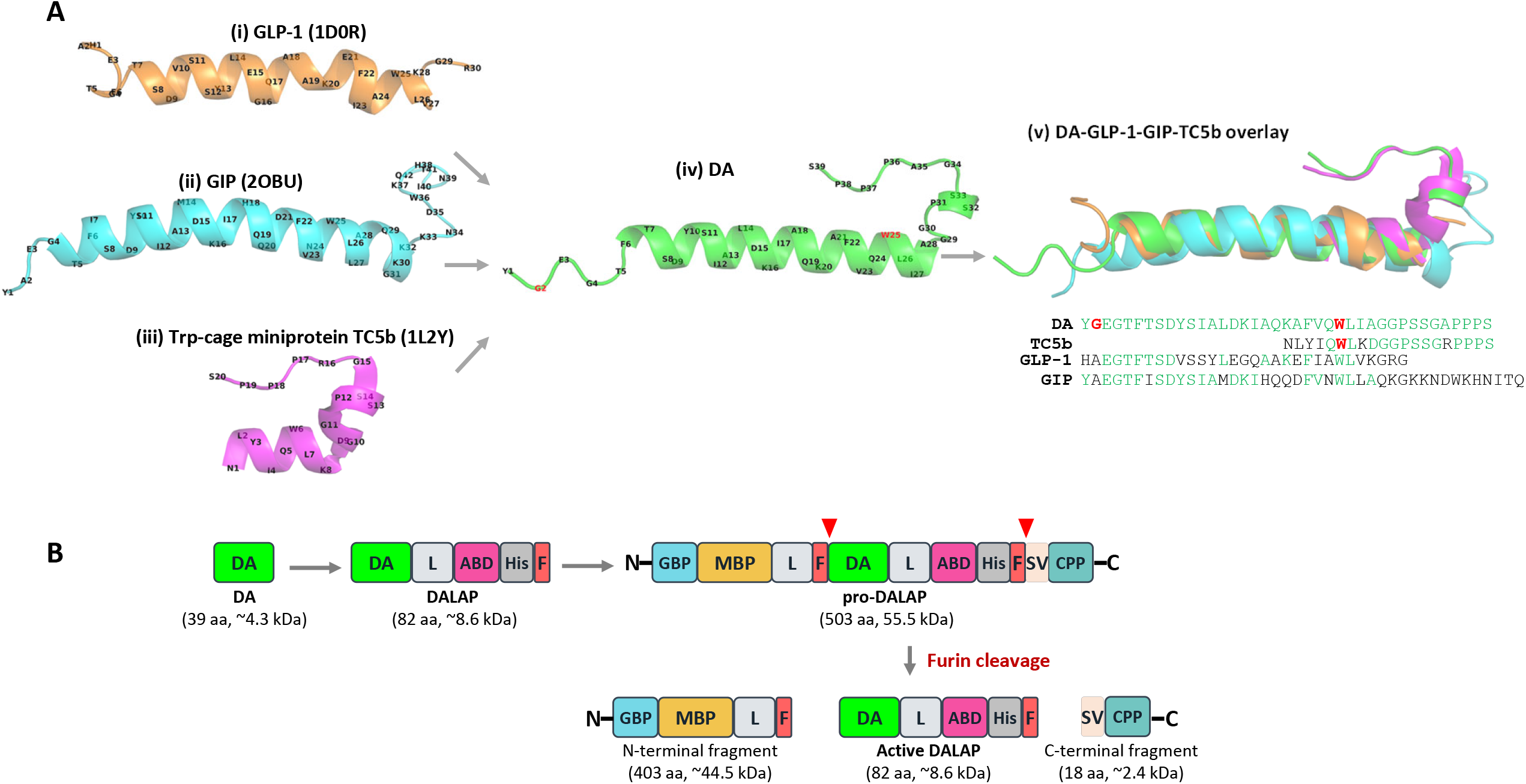
Structure-guided design of the dual agonist (DA) and modular construction of pro-DALAP. **(A)** Design and structural comparison of DA. Human glucagon-like peptide-1 (GLP-1), human glucose-dependent insulinotropic polypeptide (GIP), and the Trp-cage miniprotein TC5b served as structural references for DA design. Cartoon representations show (i) GLP-1 (PDB: 1D0R; orange), (ii) GIP (PDB: 2OBU; cyan), (iii) TC5b (PDB: 1L2Y; magenta), and (iv) the AlphaFold-predicted DA structure (green). Residue identities and positions are labeled, with Gly2 (G2) and Trp25 (W25) highlighted in red in DA. (v) Structural superposition of DA, GLP-1, GIP, and TC5b, with their corresponding amino acid sequences shown below. Residues identical to DA at the displayed sequence positions are colored green; DA Gly2 and Trp25 and the corresponding TC5b Trp6 are highlighted in red. **(B)** Schematic illustration of the stepwise design of pro-DALAP. The dual agonist peptide (DA; 39 amino acids) is extended with a linker, an albumin-binding domain, a hexa-histidine tag, and a furin cleavage site to generate DALAP (82 amino acids). Additional modules are incorporated to generate the pro-DALAP fusion precursor (503 amino acids; approximately 55.5 kDa). Furin cleavage at the sites indicated by red arrowheads produces an N-terminal fragment (403 amino acids; approximately 44.5 kDa), active DALAP (82 amino acids; approximately 8.6 kDa), and a C-terminal fragment (18 amino acids; approximately 2.4 kDa). GBP, GM1-binding peptide; MBP, maltose-binding protein; L, linker; F, furin cleavage site; DA, dual agonist; ABD, albumin-binding domain; His, hexa-histidine tag (6×His); SV, serine and valine residues; CPP, cell-penetrating peptide. N and C indicate the amino and carboxyl termini, respectively.

The substitution of Ala2 with Gly2 was intended to reduce susceptibility to dipeptidyl peptidase IV (DPP-IV)-mediated cleavage. Modification of the C-terminal region was informed by the lower evolutionary conservation of GIP residues 31–42 relative to its N-terminal and core regions [31]. This observation provided a rationale for exploring C-terminal sequence modifications while retaining much of the GIP-derived receptor-interacting sequence.

The Trp-cage miniprotein TC5b provided a structural reference for evaluating the proposed C-terminal motif. TC5b adopts a compact fold in which an α-helical segment and a folded-back C-terminal region surround a buried tryptophan side chain. Barua et al. (2008) [32] demonstrated that Trp burial, hydrophobic packing, and interactions between the helical and C-terminal regions contribute to fold stability. In DA, Trp25, Phe22, and the C-terminal proline-rich segment were intended to support analogous hydrophobic packing. These features were incorporated with the aim of stabilizing the peptide conformation and potentially reducing proteolytic susceptibility; however, formation of a Trp-cage-like motif and protection against neutral endopeptidase (NEP)-mediated degradation remain to be experimentally established.

The DA structure was predicted using AlphaFold and visualized in PyMOL alongside the experimental structures of GLP-1 (PDB: 1D0R), GIP (PDB: 2OBU), and TC5b (PDB: 1L2Y). Structural superposition and sequence comparisons were used to examine similarities between DA and these reference peptides (**Figure 1A**). Functional activation of both GLP-1 and GIP receptors by the synthetic DA peptide was subsequently demonstrated in receptor activation assays (**Figure 3A and B**).The developed peptide was further evaluated using computational protein modeling tools, including AlphaFold and PyMOL.

Although our synthetic dual agonist was engineered to exhibit an extended half-life and enhanced receptor affinity relative to endogenous incretins, it is unlikely to induce severe hypoglycemia, even under conditions of overdose. This is because the glucose-lowering actions of both GLP-1 and GIP are inherently glucose-dependent [26]. Upon binding of GLP-1 or GIP to their respective receptors on pancreatic β-cells, intracellular cAMP is generated, which amplifies insulin secretion. Notably, cAMP production is closely coupled to cellular ATP levels derived from glucose metabolism [4]. Under low-glucose conditions, reduced ATP availability limits cAMP generation, thereby attenuating downstream signaling and preventing excessive insulin release.

Once the peptide sequence was finalized and its efficacy validated, a maltose-binding protein (MBP) was fused to enhance solubility and stability during expression in engineered yeast (**Figure 1B**). To facilitate interaction with intestinal epithelial cells following oral delivery, two targeting ligands were incorporated: a GM1-binding peptide [33] and a cell-penetrating peptide derived from the human transcription factor PDX1 [34, 35] (**Figure 1B**). To further prolong systemic exposure, a human serum albumin–binding domain was included to extend circulating half-life [36] (**Figure 1B**). In addition, furin cleavage sites, RKKR, were introduced at both termini of the peptide [37, 38] to enable proteolytic conversion into the active form within the human intestine (**Figure 1B**). This engineered fusion construct was designated pro-DALAP (Dual Agonist Long-Acting Peptide). The complete amino acid sequence of pro-DALAP, with annotated functional domains and furin cleavage sites, is provided in **Supplementary Figure 1**.

### Design of the Pro-DALAP Expression Construct and Genome Integration of Edible Yeast Strain

To generate a yeast strain harboring a single copy of the pro-DALAP expression cassette, *Saccharomyces cerevisiae* S288C was employed. The *URA3* locus was first disrupted using 100 bp DNA fragments homologous to the promoter and 5′ coding regions. Successful knockout strains were selected on synthetic defined (SD) medium supplemented with uracil and 5-fluoroorotic acid (5-FOA), which is converted into a toxic compound in the presence of a functional *URA3* gene; thus, only *ura3* knockout cells survive. The verified knockout strain was subsequently used for transformation with a single-copy pro-DALAP expression cassette. For genomic integration, three codon-optimized synthetic DNA fragments (1,190 bp, 1,350 bp, and 1,686 bp) were co-transformed to facilitate homologous recombination at the disrupted *URA3* locus. These fragments contained overlapping regions that enabled in vivo assembly into a single contiguous construct, along with flanking homology arms targeting the integration site. The assembled cassette included a functional wild-type *URA3* gene, allowing selection of successful transformants on SD medium lacking uracil. To drive pro-DALAP expression, the strong constitutive triosephosphate dehydrogenase 3 (*TDH3*) promoter and the enolase 2 (*ENO2*) terminator were employed. Correct single-copy integration of the pro-DALAP cassette was confirmed by sequencing of PCR-amplified genomic DNA (**Figure 2A**).

**Figure 2.**
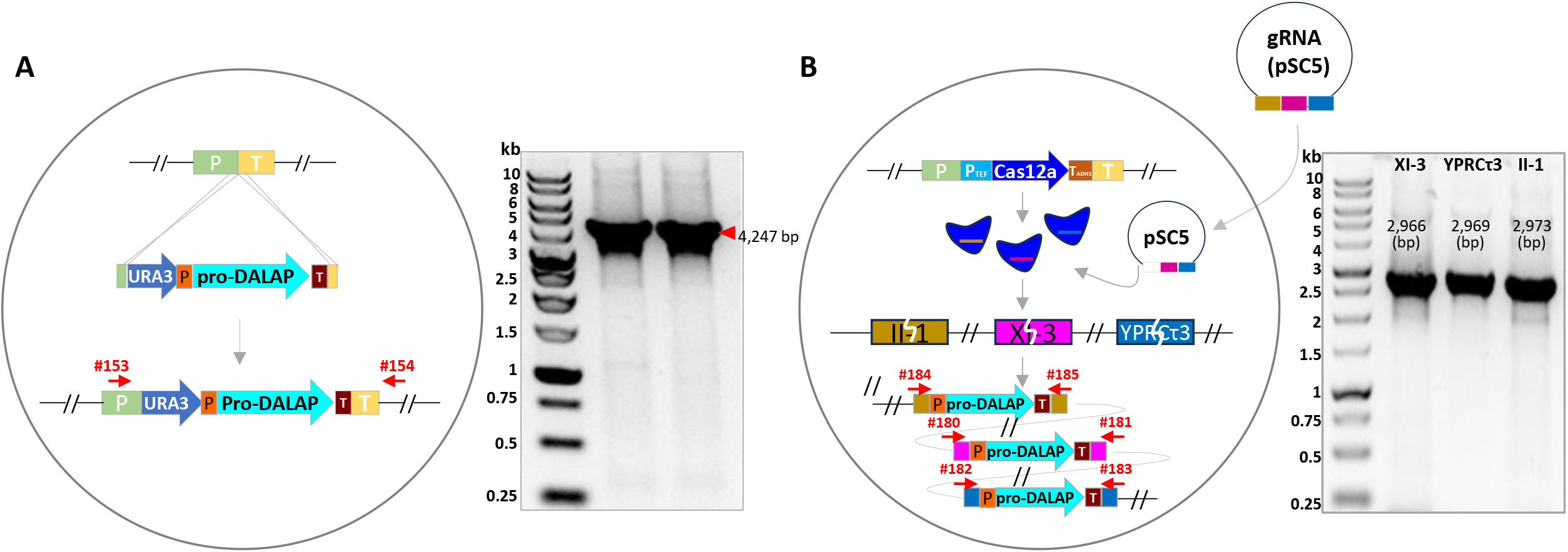
Strategies for single-copy and three-copy genomic integration of pro-DALAP expression cassettes in *Saccharomyces cerevisiae*. (A) Single-copy integration of the pro-DALAP expression cassette, driven by the *TDH3* promoter and flanked downstream by the *ENO2* terminator, at the *URA3* locus. The schematic illustrates insertion of the cassette and restoration of the *URA3* selection marker. PCR verification is shown on the right, with an expected amplicon size of 4,247 bp using primers #153 and #154. (B) CRISPR/Cas12a-mediated integration of three pro-DALAP expression cassettes. Guide RNAs expressed from plasmid pSC5 target the II-1, XI-3, and YPRCτ3 loci. Each expression cassette comprises the TDH3 promoter (P), pro-DALAP coding sequence, and ENO2 terminator (T). PCR verification is shown on the right, with expected amplicon sizes of 2,966 bp (XI-3), 2,969 bp (YPRCτ3), and 2,973 bp (II-1). Numbered red arrows indicate the positions and orientations of primers used to verify genomic integration.

To evaluate whether increased gene copy number enhances pro-DALAP expression, a yeast strain carrying three copies of the pro-DALAP gene was generated. A Cas12a expression cassette, driven by the constitutive TEF promoter and containing the ADH1 terminator, was first transiently integrated at the *URA3* locus using the strategy described for the single-copy strain [18].

Three pro-DALAP expression constructs were assembled by overlap PCR, each containing distinct left and right homology arms targeting one of three genomic loci: II-1, XI-3, and YPRCτ3. Each construct was assembled from four DNA fragments (203 bp, 1,319 bp, 1,586 bp, and 203 bp), with the two 203 bp fragments providing the locus-specific homology arms. The resulting three 2,965 bp expression cassettes were sequence-verified and co-transformed into the protease-deficient yeast strain together with the guide RNA plasmid pSC5, which carried a *URA3* selection marker. Transformants were selected on minimal SD medium, and correct integration of one pro-DALAP cassette at each target locus was confirmed by PCR and sequencing (**Figure 2B**).

Following verification, the guide RNA plasmid was eliminated by culturing cells in non-selective rich YPD medium. After plasmid loss was confirmed, the Cas12a expression cassette at the *URA3* locus was replaced with a functional *URA3* gene using the strategy described for the single-copy strain. For both the single-copy and three-copy strains, genomic DNA sequencing was performed across approximately 10 kb regions surrounding the relevant integration sites (II-1, XI-3, YPRCτ3, and *URA3*). No deleterious mutations associated with genome editing were detected in the sequenced regions.

### Expression and Physiological Characterization of Engineered Yeast

To analyze protein expression in the single-and three-copy strains, cells were cultured in YPD medium and subsequently harvested for lyophilization. The dried yeast cells were then ground using a mortar and pestle or a coffee grinder. Total protein was extracted from sonication-treated homogenates and used for Western blot and densitometry analyses. To evaluate pro-DALAP expression using an anti-MBP antibody, purified MBP protein was used to generate a linear regression curve, which was applied to interpolate band intensities obtained from the iBright 1500 imager for each strain. Six Western blot images, each loaded with protein samples from both strains, were analyzed, yielding a total of 18 quantification measurements. The mean expression levels of the single-and three-copy strains were 1.27 mg/g (± 0.21 SD, dry weight) and 4.05 mg/g (± 0.31 SD, dry weight), respectively (**Figure 3A**). Thus, the three-copy strain exhibited a 3.2-fold increase in pro-DALAP expression compared with the single-copy strain.

**Figure 3.**
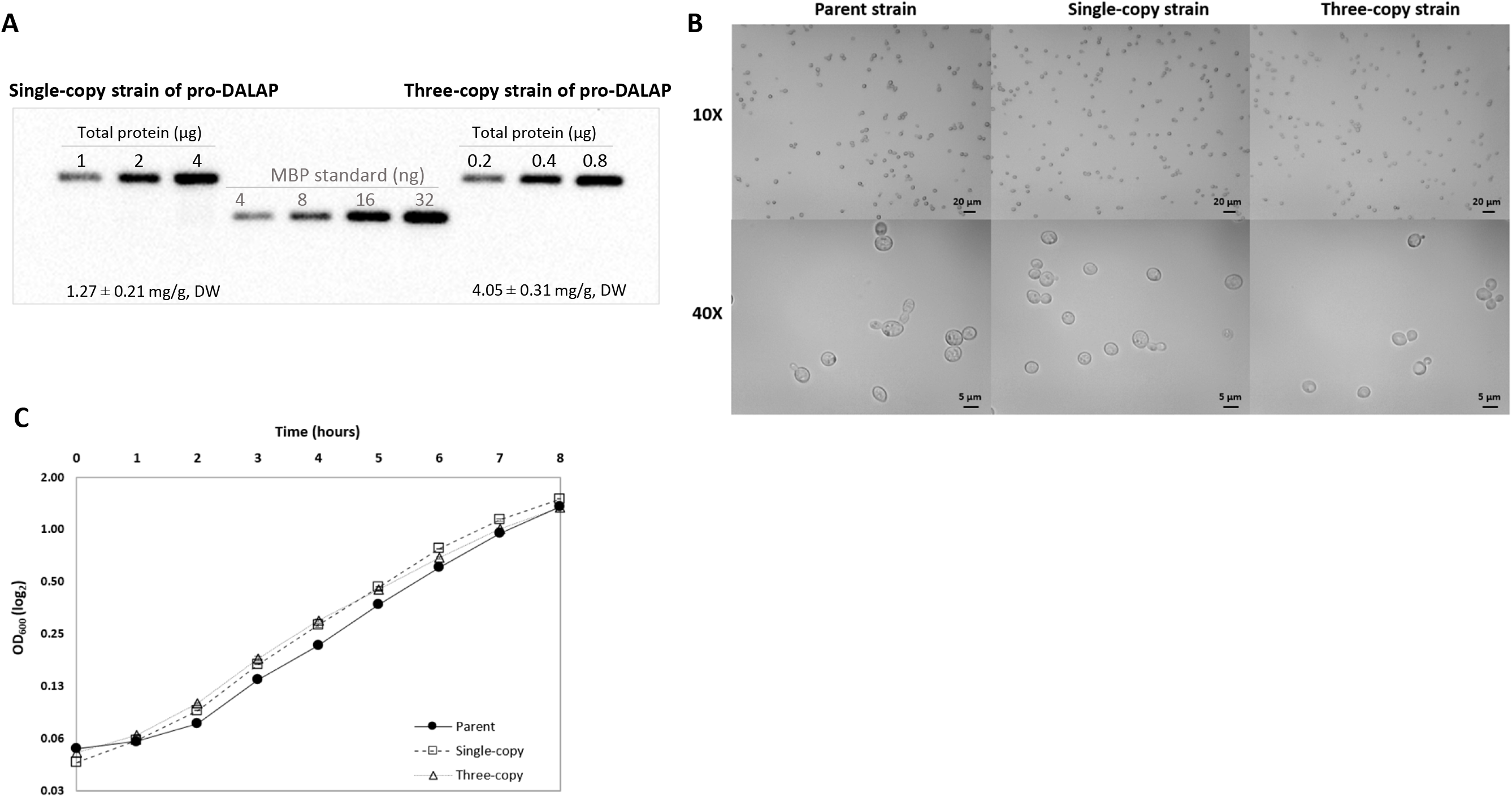
Characterization of transformed yeast expressing pro-DALAP. **(A)** Comparison of pro-DALAP expression levels in single-copy and three-copy strains. Western blot and densitometric analyses were performed to assess pro-DALAP expression in single-copy and three-copy strains. Total proteins extracted from both strains were analyzed by SDS-PAGE, with the indicated amounts of total protein loaded per lane. MBP standards (4–32 ng) were used for densitometric interpolation. Calculated pro-DALAP levels (mg/g ± SD dry weight [DW]) are shown below each gel image. **(B)** Comparison of cell morphology between parent and transformed strains. Cells were examined under a light microscope at the indicated magnifications to assess any abnormal morphology or budding characteristics. Each strain was cultured in 5 mL of YPD at 30°C with shaking at 250 rpm for 72 hours. Five microliters of undiluted culture were spotted onto glass slides for imaging. **(C)** Growth curves of parent, single-copy, and three-copy yeast strains in YPD medium. OD₆₀₀ values (log₂ scale) of the parent strain (●, solid line), single-copy strain (□, dashed line), and three-copy strain (△, dotted line) were measured hourly over an 8-hour period. Measurements were performed in triplicate for each strain. Error bars (where applicable) represent the standard error of biological replicates. Cryopreserved stocks of each strain were revived for one week and subcultured daily by inoculating 5 µL into 5 mL of YPD to ensure full recovery. Cultures were then diluted to an OD₆₀₀ of approximately 0.05, and growth was monitored by measuring OD₆₀₀ every hour.

To investigate any potential deleterious effects of pro-DALAP expression on yeast growth, we performed microscopic observations of the single- and three-copy strains alongside the parental wild-type strain and found no differences in morphology (**Figure 3B**). This suggests that the genetic modifications did not affect cell shape or budding under conditions similar to those used in commercial growth settings. We further compared the growth rates of the parental and transformant strains in YPD medium at 30 °C with shaking at 250 rpm, measuring OD₆₀₀ every hour (**Figure 3C**). Maximum specific growth rates were determined by plotting the natural logarithm of OD₆₀₀ over time and applying linear regression to the exponential growth phase using at least five data points. The untransformed parental strain exhibited a growth rate of 0.438 h⁻¹, while the single-copy and three-copy strains showed growth rates of 0.468 h⁻¹ and 0.433 h⁻¹, respectively. These results indicate no significant physiological differences among the strains, as their growth rates were nearly identical under the tested conditions.

### Functional Validation of the Dual Agonist Peptide

The biological activity of the cleaved DALAP peptide was assessed using GPCR-based assays (**Figure 4A** and **B**). GLP-1 receptor activation was measured using Exendin-4 as a positive control, in the presence or absence of human serum albumin (HSA). In the GLP-1 receptor potency assay, DALAP in the absence of human serum albumin (HSA) (RC₅₀ = −6.44) exhibited approximately 11-fold greater potency than DALAP in the presence of HSA (RC₅₀ = −5.40). In contrast, Exendin-4 showed only a modest 1.45-fold difference between conditions (RC₅₀ = −4.95 without HSA vs. −5.11 with HSA), consistent with the absence of an albumin-binding domain and the minimal influence of HSA on its activity. This limited shift contrasts with the pronounced reduction in DALAP potency observed upon HSA addition. The decreased activity of DALAP in the presence of HSA is likely attributable to interactions between its albumin-binding domain and HSA, which may transiently sequester the peptide and reduce receptor engagement. Despite this effect, DALAP (RC₅₀ = −6.44) remained approximately 30-fold more potent than Exendin-4 (RC₅₀ = −4.95) under HSA-free conditions. Notably, even in the presence of HSA, DALAP (+HSA; RC₅₀ = −5.40) retained an approximately 2-fold higher potency compared to Exendin-4 (+HSA; RC₅₀ = −5.11).

**Figure 4.**
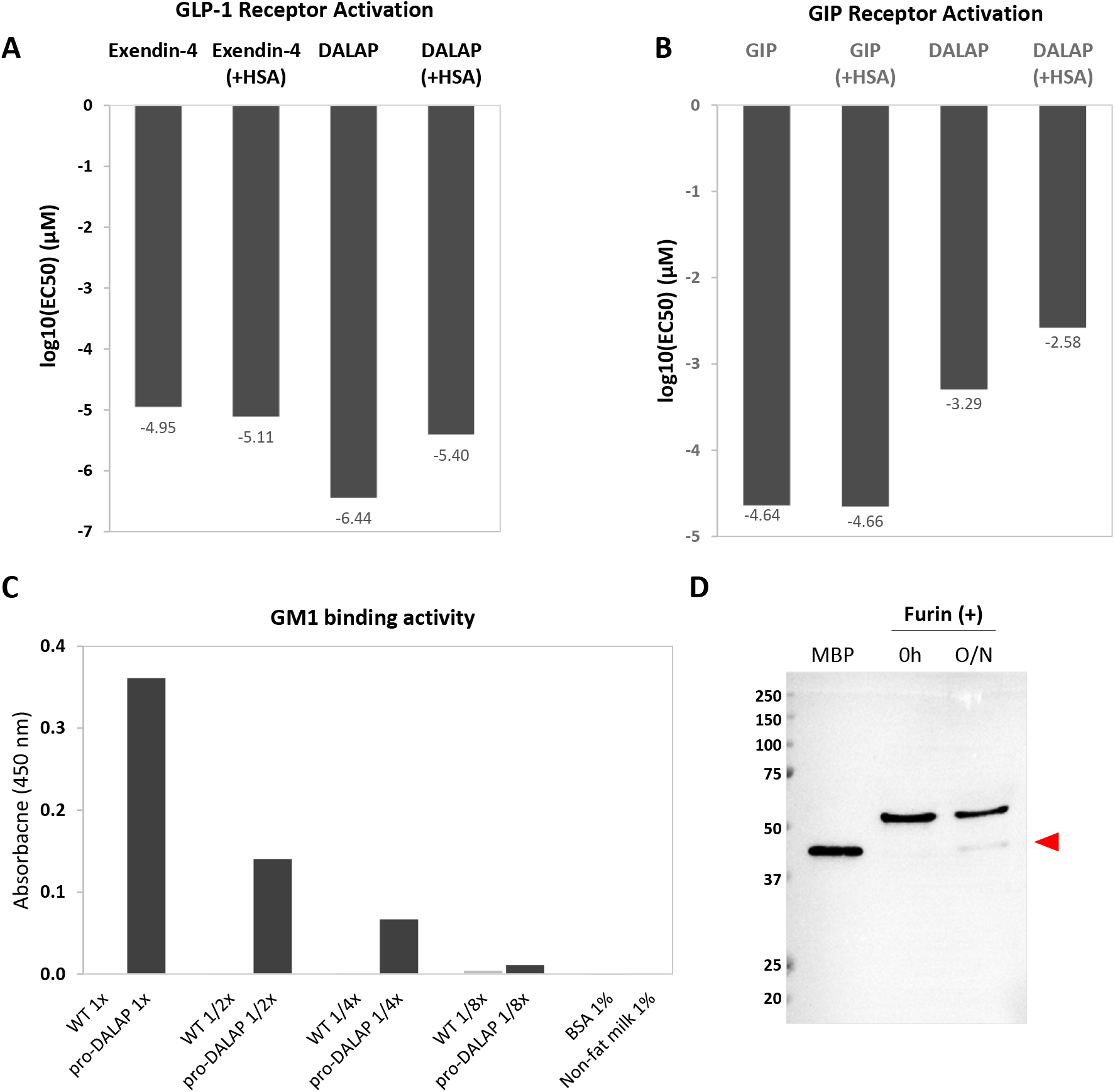
Characterization of the functional activities of DALAP and pro-DALAP. (A and. **B)** Interaction of DALAP with the GLP-1 receptor **(A)** and GIP receptor **(B)**. Binding activity of DALAP to the receptors was indirectly assessed using a G-protein–coupled receptor (GPCR) signaling assay linked to adenylate cyclase, performed in the presence or absence of human serum albumin (HSA). DALAP was incubated with HEK293 cells expressing the respective GPCR, and changes in luminescence corresponding to intracellular cAMP levels were measured as a functional readout of GPCR activation. **(C)** Interaction of the N-terminal GM1-binding peptide of pro-DALAP with GM1. One microgram of soluble protein extracted from untransformed baker’s yeast (WT) and pro-DALAP–expressing yeast was serially diluted as indicated. Diluted samples were incubated in 96-well plates coated with GM1. Binding was evaluated by ELISA, and absorbance was measured at 450 nm following TMB (3,3′,5,5′-tetramethylbenzidine dihydrochloride) development using horseradish peroxidase–conjugated secondary antibodies. WT indicates soluble protein from untransformed yeast; pro-DALAP indicates soluble protein from pro-DALAP-expressing yeast. BSA (bovine serum albumin) and non-fat milk were used as negative controls. **(D)** Proteolytic cleavage of pro-DALAP by furin. Purified pro-DALAP (55.5 kDa) was incubated with furin, and cleavage was analyzed by Western blot. The furin-cleaved product (44.5 kDa) is indicated by a red arrow. S, solubilizer (42 kDa); O/N, overnight incubation.

Next, GIP receptor activation was evaluated using GIP as a positive control, with and without HSA. In this assay, DALAP in the absence of HSA (RC50, -3.29) showed approximately 5-fold greater potency than DALAP in the presence of HSA (RC50, -2.58). By contrast, GIP displayed nearly identical potency in both conditions (RC50, -4.64 without HSA vs. -4.66 with HSA), indicating no meaningful effect of HSA. The reduction in DALAP activity under HSA conditions is again consistent with interactions involving the albumin-binding domain. Comparative analysis at the GIP receptor further showed that DALAP (RC50, -3.29) was approximately 22-fold less potent than GIP (RC50, -4.64). This difference became more pronounced in the presence of HSA, where DALAP (+HSA; RC50, -2.58) was approximately 120-fold less potent than GIP (+HSA; RC50, -4.66).

Collectively, these results demonstrate that DALAP retains strong intrinsic GLP-1 receptor activity but is highly sensitive to the presence of human serum albumin (HSA), likely due to interactions mediated by its albumin-binding domain. While HSA substantially reduces DALAP potency at both GLP-1 and GIP receptors, this effect is markedly more pronounced than that observed for reference ligands (Exendin-4 and GIP), which remain largely unaffected, likely because they lack an albumin-binding domain. Importantly, despite this reduction, DALAP maintains superior potency over Exendin-4 at the GLP-1 receptor even in the presence of HSA, supporting its potential as an effective GLP-1 agonist. However, its comparatively weak potency at the GIP receptor, further diminished under HSA conditions, suggests that its dual agonist profile may be functionally biased toward GLP-1 activity. Overall, these findings highlight both the therapeutic promise of DALAP and the critical influence of albumin binding on its pharmacological performance.

To evaluate the binding affinity of the GM1-binding peptide to GM1, a GM1-binding ELISA assay was performed. Soluble proteins extracted from both wild-type and pro-DALAP-expressing yeast strains were adjusted to 1 µg/µL, then serially diluted and incubated in GM1-coated 96-well plates. As shown in **Figure 3C**, binding activity was detected in the soluble fraction from yeast expressing pro-DALAP, with a linear decrease in signal observed upon twofold dilution, indicating a specific interaction mediated by pro-DALAP. In contrast, no binding was detected in the soluble fraction from the wild-type strain or in negative controls, including non-fat dry milk and bovine serum albumin (BSA) (**Figure 3C**).

Pro-DALAP was designed to be processed endogenously during transcytosis across epithelial cells following binding via receptor-ligand or electrostatic interactions. Accordingly, furin cleavage sites were introduced at both termini to enable conversion of pro-DALAP into the active peptide, DALAP. As shown in **Figure 3D**, furin-mediated cleavage was confirmed: incubation of the purified 55.5 kDa pro-DALAP with furin generated a cleaved product at the expected size of approximately 44.5 kDa. These results demonstrate that furin correctly recognizes the engineered cleavage sites in pro-DALAP and efficiently produces the active peptide.

### Toxicological Evaluation in Rats

Once we confirmed the binding potency of DALAP to GLP-1 and GIP receptors, the expression of pro-DALAP in yeast, and the functionality of its domains, we conducted animal studies to evaluate toxicity and safety. These studies assessed DALAP, administered via injection. In the study with invasive delivery of active DALAP peptide, a two-phase toxicology study was conducted in Sprague-Dawley rats to assess the safety profile of DALAP following intravenous administration. In phase I (dose escalation), single doses of DALAP were administered via intravenous injection at dose levels of 10, 100, and 300 µg/kg (n = 2 males per group). All animals were monitored for 5 days post-dosing and subsequently euthanized for gross pathological examination on Day 6. No mortalities or treatment-related adverse effects were observed at any dose level (**Supplementary Data 1**). There were no gross pathological findings. Based on these results, the maximum tolerated dose (MTD) for a single intravenous administration was determined to be ≥300 µg/kg. In phase II (repeat-dose study), DALAP was administered once daily via intravenous injection for 7 consecutive days at dose levels of 100 and 300 µg/kg/day, with a vehicle control group included (n = 2 males per group). Animals were observed for 1 day following the final dose and then subjected to necropsy on Day 8. Evaluations included mortality, clinical observations, body weight, food consumption, clinical pathology (hematology and clinical chemistry), organ weights, and both macroscopic and microscopic pathology. No mortalities or treatment-related adverse findings were observed throughout the study. There were no significant changes in any measured parameters, including clinical signs, body weight, food intake, hematology, clinical chemistry, or histopathology. Accordingly, the no observed adverse effect level (NOAEL) was established at 300 µg/kg/day following repeated intravenous administration for 7 consecutive days **(Supplementary Data 1)**.

### Effects of pro-DALAP Supplementation

Based on animal studies, the maximum tolerated dose (MTD) for invasive delivery of the DALAP peptide was 300 µg/kg, corresponding to 1,941 µg/kg of pro-DALAP protein. Using an assumed adult body weight of 84 kg, approximating the midpoint of the CDC-reported mean weights for U.S. men and women aged ≥20 years [39], this translates to approximately 163.0 mg of pro-DALAP per person for injection. Applying the rat-to-human equivalent dose conversion factor (dividing the rat dose by 6.2), along with a safety factor of 10 for human studies, yields a target injection dose of approximately 2.63 mg of pro-DALAP per person (163.0 mg divided by 62) [40]. Based on these calculations, the oral dose for the human study was determined to be approximately 4.74 mg of pro-DALAP per person. To estimate systemic availability, an oral bioavailability of 5% was assumed. At an oral dose of 4.74 mg of pro-DALAP, this corresponds to approximately 0.24 mg becoming systemically available, below the human dose estimate derived from animal MTD scaling. However, oral bioavailability has not been established for this formulation, and actual systemic availability could be higher or lower than this estimate. Therefore, this calculation does not establish the maximum possible systemic exposure or demonstrate safety in humans. Although 5% oral bioavailability was assumed for this calculation, bioavailability below 1% has been reported for some oral peptide formulations [13], suggesting that systemic availability below the assumed 5% is plausible. However, the bioavailability of the present formulation remains to be determined. For the human study, dried, powdered yeast cells expressing pro-DALAP were formulated into enteric-coated oral capsules for a 12-week, randomized, controlled, two-arm trial designed to evaluate effects on body weight, glycemic control, and appetite-related outcomes in adults aged ≥18 years. Participants were assigned to the intervention group, which received pro-DALAP supplementation, or the control group, which received dietary advice. The final baseline cohort comprised 98 participants, including 51 in the intervention group and 47 in the control group (**Figure 6A**). Participants in the intervention group received Evolv GLP-1 (BioDrive, Inc.), an oral formulation containing EV1-Peptide, consisting of yeast expressing pro-DALAP, at a daily dose providing approximately 4.74 mg of pro-DALAP. Participants in the control group did not receive the study product and followed standardized healthy eating recommendations based on American Diabetes Association guidelines. After providing informed consent, participants completed baseline questionnaires, underwent blood collection at a Quest Diagnostics center, and measured body weight and waist circumference at home before randomization.

Baseline participant characteristics are summarized in **Figure 6A**. Mean age was 46.92 ± 11.86 years in the intervention group and 50.64 ± 14.10 years in the control group. Mean baseline body weight was 74.52 ± 9.45 kg and 78.26 ± 11.30 kg, respectively, and mean BMI was 26.97 ± 1.47 kg/m² and 27.34 ± 2.48 kg/m², respectively. Women accounted for 86.27% of the intervention group and 68.09% of the control group.

An exploratory interim analysis evaluated percentage changes from baseline in body weight and waist circumference using measurements available at weeks 4, 6, and 8 at the interim data cutoff (**Figure 5A–C**). For body weight, the intervention and control groups contributed 25 and 20 measurements at week 4, 13 and 16 at week 6, and eight per group at week 8. Mean percentage changes were −0.79%, −1.19%, and −2.29% in the intervention group, compared with −0.41%, −0.19%, and +0.14% in the control group, respectively (**Figure 5A and B**). At week 8, mean changes were −2.29 ± 4.20% versus +0.14 ± 1.64% (mean ± SD), corresponding to a descriptive between-group difference of −2.43 percentage points (p = 0.162).

**Figure 5.**
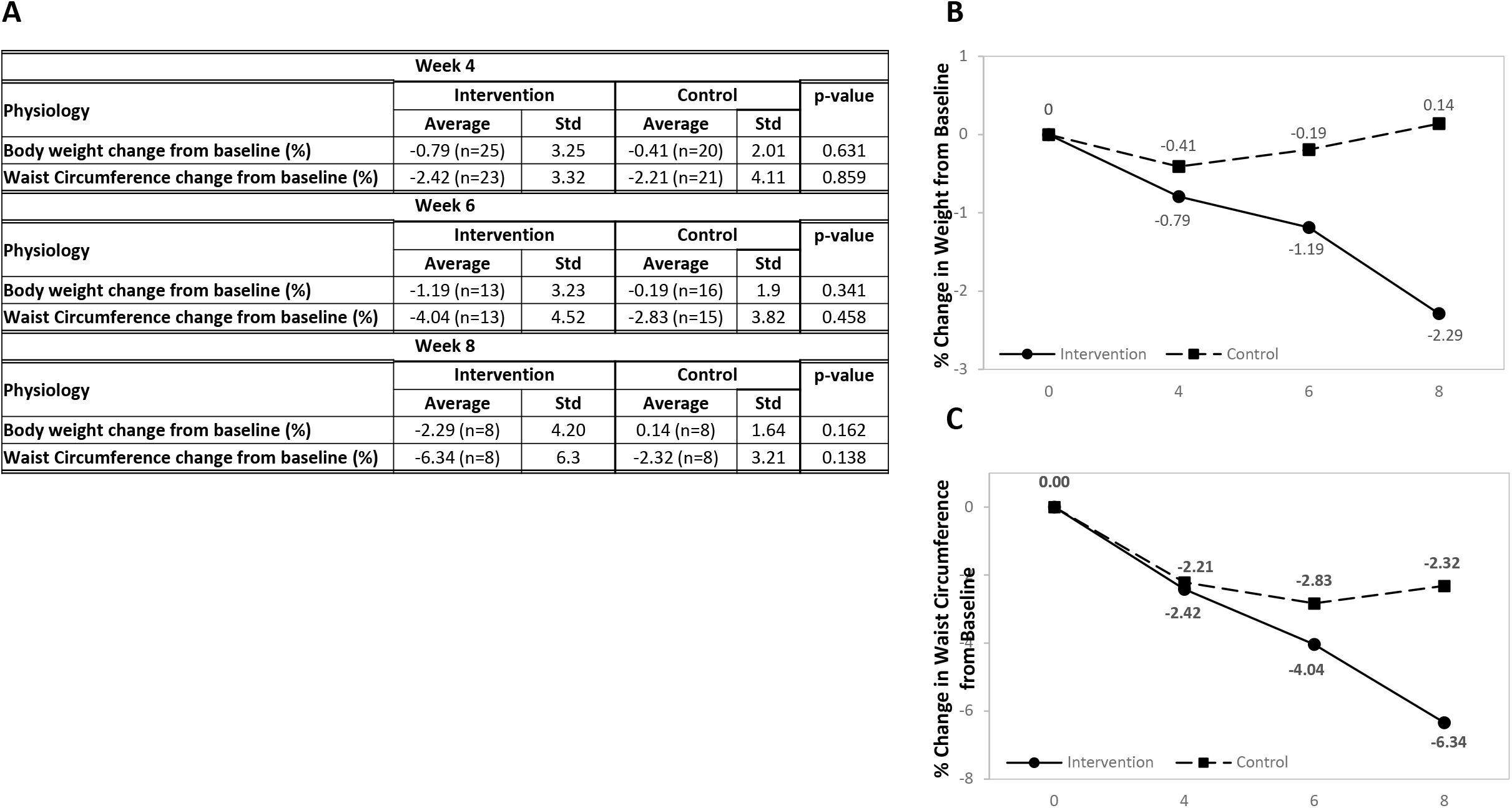
Interim changes in body weight and waist circumference during the 12-week randomized controlled trial. **(A)** Mean percentage changes from baseline in body weight and waist circumference at weeks 4, 6, and 8 in the intervention and control groups. Values are presented as means and standard deviations, with the number of participants contributing data indicated for each outcome and visit. Between-group p-values were calculated using two-sided Welch’s independent-samples t-tests. **(B–C)** Mean percentage changes in body weight **(B)** and waist circumference **(C)**. Solid lines with circles represent the intervention group; dashed lines with squares represent the control group. Baseline percentage changes are zero by definition. The interim analysis includes data available at the interim cutoff, with participant numbers varying across visits; connecting lines therefore do not necessarily represent changes in the same participants. No between-group difference was statistically significant.

For waist circumference, the intervention and control groups contributed 23 and 21 measurements at week 4, 13 and 15 at week 6, and eight per group at week 8. Mean percentage changes were −2.42%, −4.04%, and −6.34% in the intervention group, compared with −2.21%, −2.83%, and −2.32% in the control group, respectively (**Figure 5A and C**). At week 8, mean changes were −6.34 ± 6.30% versus −2.32 ± 3.21%, corresponding to a descriptive between-group difference of −4.02 percentage points (p = 0.138).

The interim analysis showed encouraging numerical patterns, with greater mean reductions in both body weight and waist circumference in the intervention group than in the control group (**Figure 5A–C**). These preliminary observations provide a basis for further investigation of the formulation’s potential effects, although between-group differences did not reach statistical significance. The small samples, including eight participants per group at week 8, limited the precision of the estimates, and participant availability varied across visits.

In the interim week-8 dataset, individual reductions of up to 6.4 kg (14.1 lb) in body weight and approximately 11.9 cm (4.7 inches) in waist circumference were observed in the intervention group. These values represent the maximum observed reductions for each outcome rather than average treatment responses and do not establish a treatment effect relative to the control group. For descriptive context, applying a simplified energy-equivalence assumption of 3,500 kcal per pound to the largest observed weight reduction yields approximately 880 kcal/day over 56 days. This represents a rough energy-equivalent estimate of the observed weight change, rather than measured energy intake or expenditure, and should not be interpreted as a reduction in daily calorie consumption.

To assess whether the preliminary patterns were also evident in the larger dataset, the completed analysis incorporated additional measurements at weeks 4, 6, and 8 and follow-up through week 12 (**Figure 6B–D**). The numbers of participants contributing measurements at these visits were 45, 42, 39, and 42 in the intervention group and 30, 31, 27, and 29 in the control group, respectively.

**Figure 6.**
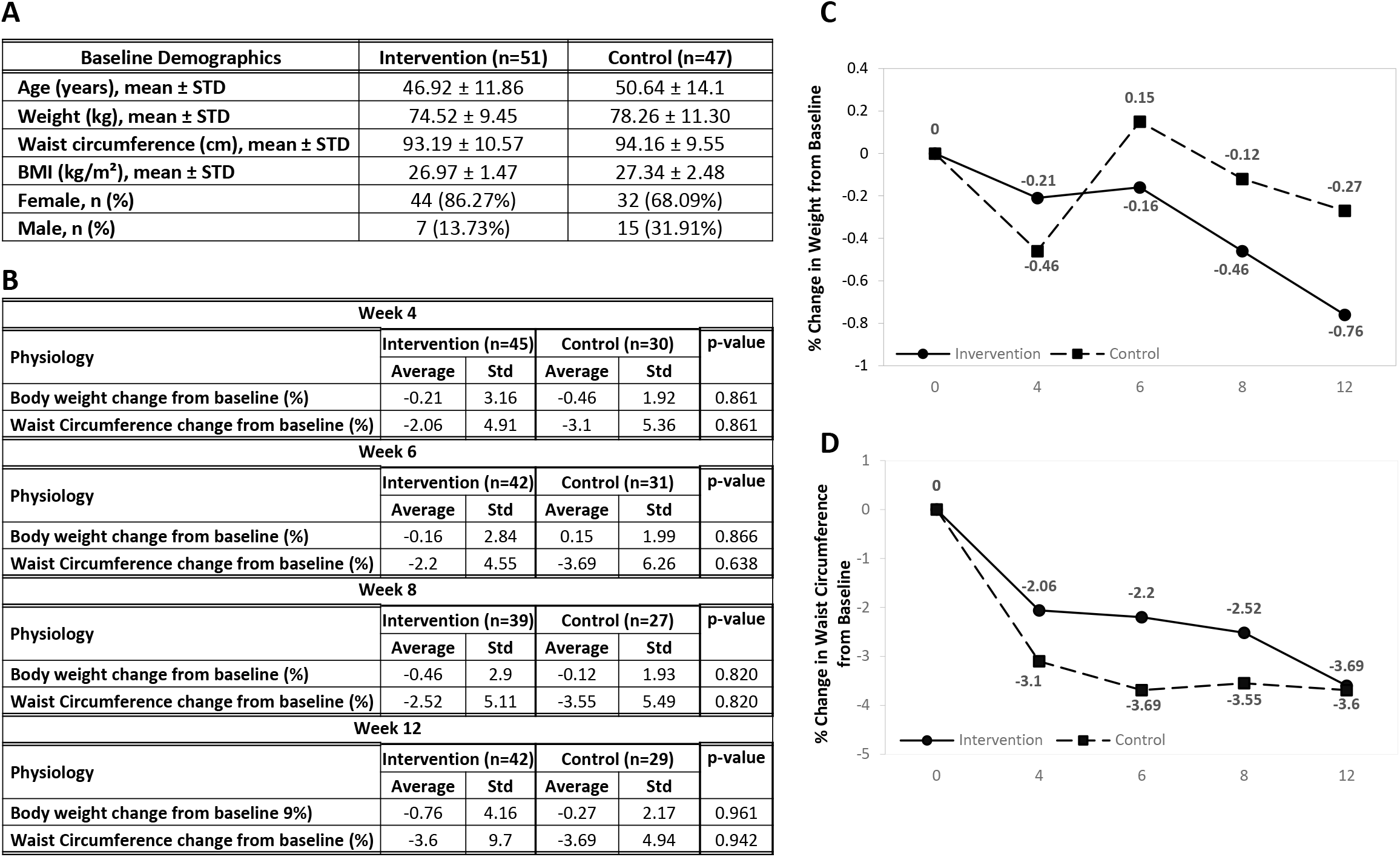
Baseline participant characteristics and percentage changes in body weight and waist circumference in the completed 12-week randomized controlled trial. (A) Baseline characteristics of participants in the intervention (n = 51) and control (n = 47) groups. Continuous variables are presented as mean ± standard deviation (SD), and sex is presented as n (%). (B) Percentage changes from baseline in body weight and waist circumference at weeks 4, 6, 8, and 12, with group-specific sample sizes, means, SDs, and p-values for between-group comparisons. (C–D) Mean percentage changes from baseline in body weight (C) and waist circumference (D). Solid lines with circles represent the intervention group; dashed lines with squares represent the control group. Baseline percentage changes are zero by definition. Sample sizes vary across visits, and connecting lines do not necessarily represent changes in the same participants. No between-group comparison was statistically significant.

Mean percentage changes in body weight from baseline were −0.21%, −0.16%, −0.46%, and −0.76% in the intervention group at weeks 4, 6, 8, and 12, respectively, compared with −0.46%, +0.15%, −0.12%, and −0.27% in the control group (**Figure 6B and C**). At week 12, the intervention group showed a numerically greater mean reduction, corresponding to a between-group difference of −0.49 percentage points. However, no between-group difference in body weight change was statistically significant at any assessed visit (all p ≥ 0.820).

Mean percentage changes in waist circumference were −2.06%, −2.20%, −2.52%, and −3.60% in the intervention group, compared with −3.10%, −3.69%, −3.55%, and −3.69% in the control group at the corresponding visits (**Figure 6B and D**). Both groups showed reductions throughout follow-up, with similar mean changes at week 12. No between-group difference in waist circumference change was statistically significant at any assessed visit (all p ≥ 0.638). Thus, the larger numerical differences favoring the intervention group in the interim subset were not reproduced in the completed analysis. Potential explanations for this difference, including variation in participant composition, adherence, and follow-up availability, are considered in greater detail in the Discussion.

Changes in HbA1c did not differ significantly between the intervention and control groups at week 6 (p = 0.999). At week 12, the control group showed a greater reduction than the intervention group (p = 0.0486) (**Supplementary Figure 2**). However, among participants with abnormal HbA1c at baseline, the proportion who achieved normal HbA1c levels was numerically higher in the intervention group than in the control group (40% vs. 29%, respectively) (**Supplementary Table 2**). Because participants were generally healthy and most had well-maintained glycemic control at baseline, there may have been limited scope for further improvement with supplementation.

Participant-reported outcomes were collected using an 18-item questionnaire (**Supplementary Data 2**), with selected positive responses summarized in **Figure 7A**. At week 4, 50.00% of respondents in the intervention group reported fewer unwanted cravings (n = 40). At week 6, 52.63% reported that their muscle definition had remained unchanged or become more noticeable (n = 38). Among week-12 respondents (n = 44), 52.27% reported improved overall well-being, and 61.36% reported unchanged or more noticeable muscle definition. In addition, 75.00% expressed a desire to continue using the product, 59.09% would recommend it to family and friends, and 56.82% indicated that they were satisfied or very satisfied with the product. Percentages were calculated using the number of respondents at each corresponding visit.

**Figure 7.**
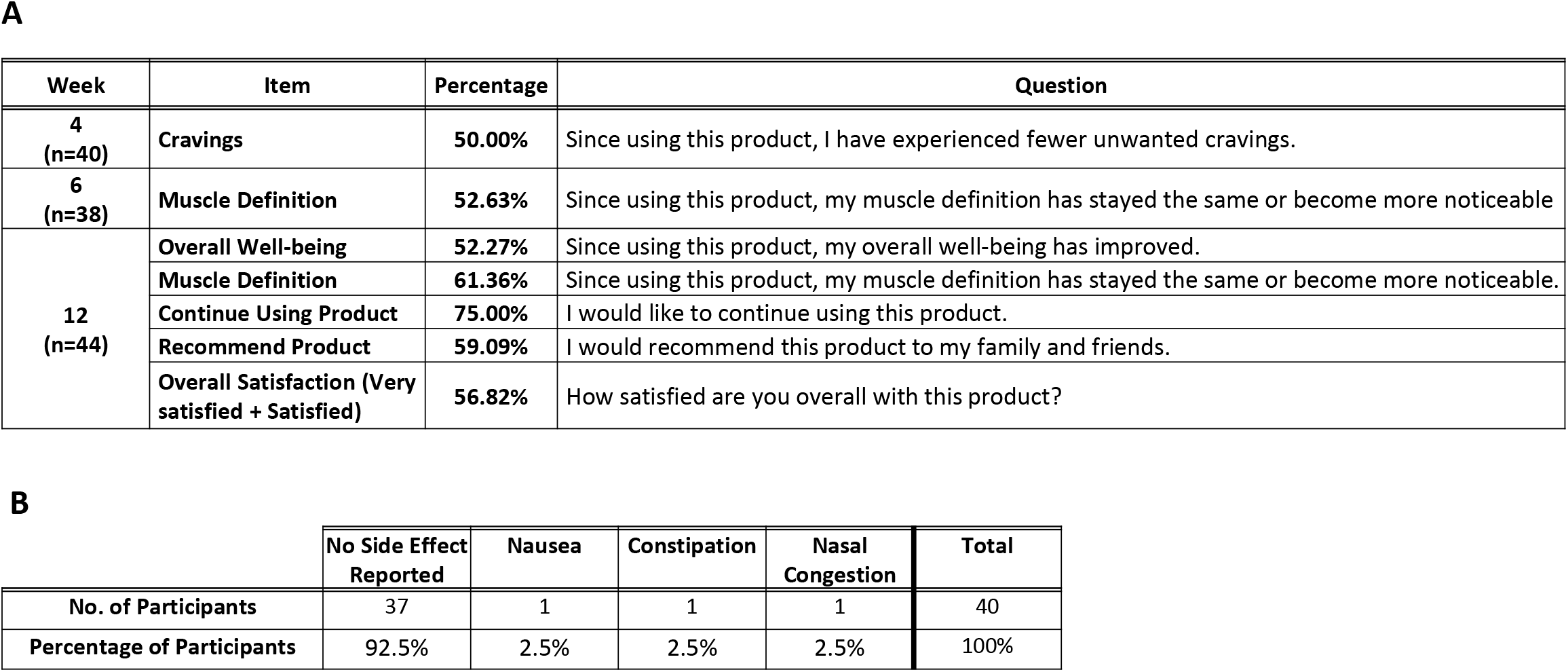
Participant-reported positive responses and side effects associated with use of EV-peptide containing pro-DALAP-expressing yeast. **(A)** Positive survey responses among participants in the intervention group at weeks 4 (n = 40), 6 (n = 38), and 12 (n = 44). Responses included reduced unwanted cravings, maintained or more noticeable muscle definition, improved overall well-being, willingness to continue using or recommend the product, and overall satisfaction. Percentages are calculated using the number of respondents at each time point. Overall satisfaction combines “very satisfied” and “satisfied” responses. **(B)** Self-reported side effects among 40 respondents out of 45 participants surveyed. Of these respondents, 37 (92.5%) reported no side effects; nausea, constipation, and nasal congestion were each reported by one respondent (2.5%). Percentages are based on the 40 respondents.

Self-reported side effects are summarized in **Figure 7B**. Of 45 participants surveyed, 40 provided responses. Among these respondents, 37 (92.5%) reported no side effects. Nausea, constipation, and nasal congestion were each reported by one respondent (2.5%) (**Supplementary Data 3**). These findings describe reported symptoms among survey respondents; they do not establish whether the symptoms were attributable to the intervention or provide a comprehensive assessment of safety.

Overall, the intervention group showed a modest numerical reduction in body weight over 12 weeks, while waist circumference decreased in both groups. The between-group comparisons did not demonstrate statistically significant effects on either measure. Selected survey responses indicated perceived benefits and product satisfaction among some intervention-group respondents. However, these subjective findings should be interpreted cautiously because participants knew whether they received the product, corresponding control-group survey results are not presented, and response rates varied across visits. Further studies with blinded outcome assessment, more complete follow-up, and prespecified analyses are needed to determine whether these perceived benefits correspond to reproducible effects on appetite or metabolic outcomes.

## Discussion

We developed a novel yeast-based platform for oral delivery of a dual agonist peptide, integrating multiple functional domains to overcome key barriers in oral biologic delivery. The pro-DALAP construct enables enhanced stability, intestinal transport, proteolytic activation, and systemic persistence.

The engineered strain used in this study is derived from *S. cerevisiae* S288C, a widely used laboratory and industrial strain, and differs only by the insertion of genes encoding the pro-DALAP. Importantly, the final product consists of non-viable, lysed yeast biomass, thereby eliminating any potential risks associated with live genetically modified organisms. The genetic modification is well-defined and limited in scope, and does not introduce traits associated with increased virulence, toxicity, or environmental persistence.

To engineer the yeast strain described here, we employed the Cas12a CRISPR system for multiplex genome editing, enabling the integration of three copies of the pro-DALAP expression cassette into targeted genomic loci in a single transformation event. Cas12a, guided by CRISPR RNA (crRNA), introduced site-specific double-strand breaks (DSBs), thereby facilitating double homologous recombination at the intended loci. Precise integration was achieved using 60-bp homologous flanking sequences on each side of the expression cassette, which were sufficient to direct insertion into the corresponding genomic regions.

To minimize the risk of off-target mutations arising from nonspecific crRNA binding and unintended DSB formation, we followed established design principles from prior studies. Specifically, insertion sites II-1 [41], XI-3 [42], and YPRCτ3 [43] were selected based on validated genomic loci. Corresponding crRNAs were designed using stringent criteria: (i) candidate sequences were ranked using the CHOPCHOP tool based on predicted activity; (ii) canonical 5′-TTTV-3′ PAM sites were identified and evaluated for RNA secondary structure using RNAfold; and (iii) sequences were screened to ensure that no secondary structures interfered with the direct repeat (DR) region, with off-target potential assessed using BLAST. The top-ranked candidates with no predicted off-target effects were selected.

To validate genomic integrity, we performed sequencing analysis covering approximately 10 kb surrounding each of the four insertion sites (II-1, XI-3, YPRCτ3, and *URA3*). A four-nucleotide deletion (ATAT) was identified at positions 1608–1611 downstream of the *URA3* stop codon (TAA). Notably, both the integration of the Cas12a gene and its subsequent replacement with *URA3* were performed without employing the Cas12a CRISPR system. Therefore, this deletion is likely to have arisen spontaneously. In addition, as shown in **Figures 3B** and **2C**, no observable differences in growth rate or cellular morphology were detected between the wild-type parental strain and the engineered strains. Collectively, these results indicate that the engineered yeast strain does not harbor deleterious mutations associated with the genome editing process.

Taken together, the extensive history of safe human consumption of *S. cerevisiae*, its strong regulatory acceptance [16, 19, 20, 22], and the controlled nature of the genetic modification support the conclusion that the engineered yeast-derived product is substantially equivalent to its parental strain. The absence of viable cells, together with the lack of detectable toxic risks and the absence of deleterious mutations, indicates that the product presents negligible safety concerns for its intended use.

Oral biologic delivery is highly desirable due to its convenience, elimination of needle-based administration, and independence from cold-chain logistics, all of which contribute to improved adherence and reduced costs [3, 35]. However, achieving systemic exposure of peptide and protein biologics via the oral route remains a major challenge. In the gastric environment, proteins and peptides undergo denaturation under highly acidic conditions and are rapidly degraded by pepsin [3]. Those that escape gastric degradation are further exposed to extensive proteolysis in the intestine by enzymes including chymotrypsin, trypsin, elastase, carboxypeptidase, and aminopeptidase [3]. To address acid-mediated degradation in the stomach, enteric coating strategies have been widely employed [3]. We validated the performance of the enteric capsules used in this study, demonstrating that they remain intact in simulated gastric conditions (pH 1.2) for up to two hours, while rapidly dissolving within 20 minutes under intestinal conditions (pH 6.8) (data not shown). Beyond physicochemical protection, we engineered DALAP to enhance epithelial interaction and promote transcytosis into systemic circulation. This was achieved through the incorporation of dual affinity modules at the N-and C-termini -a GM1-binding peptide and a cell-penetrating peptide -facilitating rapid epithelial surface binding and cellular uptake (**Figure 1B**).

GM1 is a glycosphingolipid enriched in lipid raft domains and typically constitutes ∼0.1 -1% of total plasma membrane lipids, corresponding to an estimated surface density on the order of 10⁶ -10⁷ molecules per cell [44–46]. This provides a high-capacity platform for ligand binding and transcytosis. GM1 is continuously present and relatively stable on the apical membrane and is actively involved in endocytic and transcytotic pathways [47]. Accordingly, when ligands bind to GM1, the resulting complexes can be internalized within minutes to hours and subsequently recycled back to the membrane or trafficked intracellularly [47]. In addition, the surface area of the small intestine is approximately 30 m² [48]. Given the high number of GM1 molecules per epithelial cell and the extensive surface area, the small intestine represents an ideal site for oral delivery of macromolecules. Furthermore, its highly convoluted structure and considerable length (approximately 6 meters) [49] provide sufficient residence time for orally delivered molecules to interact with the epithelium and be absorbed.

Although we confirmed strong interaction between the GM1-binding peptide and GM1 using a GM1 ELISA assay (**Figure 4C**), we further sought to enhance the attachment of pro-DALAP to epithelial cells and promote subsequent absorption. To this end, we incorporated a cell-penetrating peptide (CPP) at the C-terminus (**Figure 1B**), which is capable of delivering attached molecules into living cells without requiring specific receptor interactions [34, 35, 50]. CPPs are typically short cationic peptides, 8–16 amino acids in length, and have demonstrated high delivery efficiency for biologically active proteins in numerous in vitro and in vivo studies using cultured mammalian cells and animal models [34, 35]. In this study, we employed a 16-amino-acid CPP sequence (RHIKIWFQNRRMKWKK) derived from human pancreatic and duodenal homeobox factor-1 (PDX-1), a transcription factor involved in pancreatic development and regulation of insulin gene expression [34, 50]. Notably, an oral delivery study using powdered, lyophilized lettuce cells expressing GFP–PTD (with the PTD comprising the same amino acid sequence as used in this study) in mice demonstrated accumulation of GFP signals in multiple organs, including the liver and lungs, as well as detectable levels in serum [35]. This study further showed that GFP fused to PTD exhibited a comparable biodistribution to that of other absorption tags, such as the non-toxic cholera toxin B subunit (CTB–GFP) and the dendritic cell–targeting peptide (DCpep–GFP) [35].

Although DALAP was engineered to extend its half-life in circulation by introducing a glycine at the second position to confer resistance to DPP-IV-mediated degradation and by incorporating a tryptophan cage (a C-terminal bent structure formed by a tryptophan at position 25, Trp cage) to protect the peptide from degradation by neutral endopeptidase (NEP) [30, 32], its direct release from an enteric capsule into the intestinal lumen may still be detrimental due to enzymatic degradation by a variety of digestive proteases [3]. Because the first two amino acids of DALAP are critical for receptor binding and subsequent signal activation, protection of the N-terminus from proteases, including aminopeptidases, is essential. Likewise, protection of the C-terminus from peptidases such as carboxypeptidases is critical, particularly because this region was engineered to extend the half-life of DALAP through interaction with human serum albumin via incorporation of an albumin-binding domain (**Figure 1B**). Accordingly, N-and C-terminal affinity tags incorporated into DALAP serve dual functions: shielding the peptide from proteolytic degradation and facilitating attachment to epithelial cells. In addition, the N-terminal bulky maltose-binding protein (MBP) plays an important protective role in the intestine. MBP not only enhances the solubility of pro-DALAP during expression in yeast but also provides steric protection against intestinal proteases such as DPP-IV and NEP. Some peptide-degrading (oligo)peptidases preferentially act on substrates of approximately 3– 100 amino acids [51]. This substrate preference provides a rationale for investigating fusion-mediated protection but does not establish resistance of pro-DALAP to intestinal proteolysis.

Once pro-DALAP reaches the intestine, the inactive precursor must be converted into its active form, DALAP. Many proteins and peptides are synthesized as inactive precursors and subsequently processed by the subtilisin/kexin-like proprotein convertase (PC) family, a group of endoproteolytic enzymes that recognize single or paired basic amino acid residues [37]. PCs are involved in the activation of a wide range of substrates, including peptide hormones, neuropeptides, growth and differentiation factors, adhesion molecules, receptors, coagulation factors, plasma proteins, extracellular matrix proteins, and proteases, as well as exogenous proteins such as viral glycoproteins (e.g., HIV-1 and influenza) and bacterial toxins (e.g., diphtheria and anthrax toxins) [37]. Among the seven PC family members in mammals, furin is the best characterized; it is ubiquitously expressed and has been implicated in proprotein processing associated with various diseases, including Alzheimer’s disease, cancer, anthrax, and Ebola [37]. Accordingly, we leveraged the endogenous proprotein activation pathway by introducing a furin cleavage site (RKKR) at both termini of DALAP. The furin recognition sequence was selected based on previous studies analyzing cleavage sites across proteins from mammals, bacteria, and viruses [38]. In this design, the second furin cleavage site in pro-DALAP is followed by serine and valine (SV), which have been identified as among the most frequently occurring residues immediately downstream of furin cleavage sites. As shown in **Figure 4D**, these cleavage sites were properly recognized and efficiently processed by furin. It is therefore assumed that DALAP, generated through endogenous proteolytic activation of pro-DALAP during epithelial transcytosis, reaches systemic exposure.

As described above, affinity tags were incorporated at both termini of pro-DALAP to promote intestinal epithelial uptake, and an albumin-binding domain was included with the intention of extending systemic exposure. In a previous oral delivery study using lyophilized lettuce expressing GFP fused to the non-toxic, GM1-binding cholera toxin B subunit (CTB), a PDX1-derived protein transduction domain (PTD), or a dendritic cell–specific binding peptide (DCpep), the percentages of GFP detected in mouse serum were 0.024%, 0.0072%, and 0.041%, respectively, based on our calculations using the data in the paper [35]. These findings illustrate the potential for low systemic delivery through such approaches, although differences in the expressed protein, delivery matrix, and experimental conditions preclude direct estimation of DALAP bioavailability from these values. Accordingly, the amount of active DALAP reaching systemic circulation following oral administration remains to be determined.

Although DALAP exhibits potent GLP-1 receptor activation in vitro, this activity alone does not establish that oral administration achieves sufficient exposure for clinically meaningful effects. For therapeutic development, purification or enrichment of the peptide could increase the administered peptide dose while reducing the amount of yeast biomass required; however, the resulting systemic exposure and efficacy would require direct evaluation. The albumin-binding domain offers a potential strategy for prolonging circulation, with a terminal half-life of approximately 1.3 days reported in a previous study using this approach [36]. This value should not be interpreted as an established half-life for orally administered DALAP, which remains subject to pharmacokinetic characterization.

Together, the molecular design and in vitro findings support a proposed model of oral pro-DALAP delivery and action involving intestinal epithelial targeting and uptake, furin-mediated release of active DALAP, association with circulating albumin, and subsequent activation of GLP-1 and GIP receptors (**Supplementary Figure 3**). Intestinal transcytosis, systemic exposure, and prolonged circulating half-life following oral administration remain to be established in vivo.

The clinical findings should be interpreted alongside, but distinguished from, the receptor pharmacology of DALAP. The 12-week randomized controlled study evaluated the yeast-derived pro-DALAP supplement in generally healthy adults with overweight. At week 12, mean body weight decreased by 0.76% in the intervention group and 0.27% in the control group, while mean waist circumference decreased by 3.60% and 3.69%, respectively. Neither outcome differed significantly between groups (p = 0.961 for body weight and p = 0.942 for waist circumference; **Figure 4**). These findings indicate that the modest numerical reduction in body weight in the intervention group cannot be attributed conclusively to supplementation. Although the control group showed a greater reduction in HbA1c at week 12 (**Supplementary Figure 2**), the normalization rate among participants with abnormal baseline HbA1c was numerically higher in the intervention group (40% vs. 29%) (**Supplementary Table 2**). Overall HbA1c changes occurred within the normal reference range, and their clinical significance remains uncertain. Relatively well-maintained baseline glycemic control may have limited the opportunity to detect additional improvement.

The receptor assays provide a biological rationale for investigating DALAP but do not establish the mechanism underlying the clinical observations. DALAP activated both GLP-1 and GIP receptors and exhibited approximately 30-fold greater potency than Exendin-4 under albumin-free conditions and approximately twofold greater potency in the presence of human serum albumin (HSA; **Figure 4A and B**). Its potency at the GIP receptor was lower than that of native GIP, particularly in the presence of HSA. These results suggest a more favorable potency profile relative to the reference agonist at the GLP-1 receptor, although the contributions of either receptor to potential clinical effects remain unresolved. Because the assays measured cAMP responses without evaluating β-arrestin recruitment, receptor internalization, or desensitization, they do not establish signaling bias or reduced receptor desensitization. HSA also reduced DALAP’s apparent potency at both receptors, consistent with albumin interaction; however, the extent to which this interaction affects systemic exposure following oral administration remains to be determined, as discussed above.

Participant questionnaires provided additional information on perceived benefits and tolerability (**Figure 6**; **Supplementary Data 2** and **3**). Some intervention-group respondents reported fewer unwanted cravings, improved well-being, and satisfaction with the product. However, these observations do not establish effects on appetite or metabolic function because the study was open-label, corresponding control-group questionnaire results were not presented, and response rates varied across visits. Of the 40 participants who answered the side-effect questionnaire, 37 reported no side effects, while nausea, constipation, and nasal congestion were each reported by one respondent. These findings suggest that most respondents tolerated the supplement over the study period, but they do not establish symptom causality or provide a comprehensive assessment of safety. Differences in study populations, treatment regimens, follow-up, and adverse-event ascertainment also preclude conclusions about superior tolerability relative to injectable incretin therapies.

Several factors may help explain the larger numerical differences favoring the intervention group in the interim analysis compared with the completed analysis. At the interim week-8 cutoff, mean changes in body weight were −2.29% in the intervention group and +0.14% in the control group, while corresponding changes in waist circumference were −6.34% and −2.32% (**Figure 5**). These observations were based on only eight participants per group, and neither between-group comparison reached statistical significance. The completed analysis included more participants at weeks 4, 6, and 8, along with follow-up data through week 12 (**Figure 6**). Adding participants with different responses may have changed the group averages. Therefore, the smaller differences between groups in the completed analysis do not necessarily mean that the initial participants lost their early improvements.

Baseline imbalances provide another possible explanation. In the completed baseline cohort, the control group was, on average, 3.74 kg heavier than the intervention group (78.26 vs. 74.52 kg), had a slightly larger waist circumference (94.16 vs. 93.19 cm), and had a higher mean BMI (27.34 vs. 26.97 kg/m²). The intervention group also included a higher proportion of women (86.3% vs. 68.1%), and the groups differed in age distribution. These characteristics may influence weight-management responses and could have affected the comparisons as enrollment progressed. The intervention group started with a lower average body weight and BMI, which may have left less room for weight reduction. However, this does not prove that these participants had more difficulty losing weight. Comparing percentage changes also does not fully account for differences in starting weight, age, and sex. To determine whether the groups became more different as recruitment continued, the baseline characteristics of early and later participants would need to be compared. An analysis accounting for these characteristics could help assess whether they contributed to the smaller between-group differences in the completed analysis.

Differences in engagement and adherence between earlier and later participants may also have contributed. Earlier enrollees may have been particularly motivated to follow the study regimen or modify their health behaviors, potentially influencing the interim observations. However, motivation and enrollment-related differences in adherence were not established, so this explanation remains a hypothesis. The study also extended through the holiday season, when social gatherings and disruptions to usual routines may have affected dietary intake, physical activity, and adherence. Some participants reported that DALAP helped prevent weight gain and excessive snacking during this period, suggesting a perceived benefit in managing holiday-related eating behaviors. However, these comments were subjective, and holiday-related influences were not directly measured. Differences in menopausal or hormonal status could likewise have influenced responses but were not assessed.

The open-label design introduces further uncertainty because knowledge of group assignment may have influenced eating behavior, adherence, and reporting of perceived benefits. Only the control group received standardized dietary guidance; therefore, the comparison reflected differences in both pro-DALAP supplementation and dietary support. Dietary guidance may have contributed to improvements in the control group, but its contribution cannot be separated from other behavioral and participant-related factors. The generally healthy study population and relatively well-maintained baseline glycemic control may also have limited opportunities for further glycemic improvement. Although HbA1c reduction favored the control group at week 12, the observed changes occurred within the normal reference range, and their clinical significance remains uncertain.

Home measurement of body weight and waist circumference introduced additional variability. The correlation between percentage changes in these outcomes was weaker in the control group than in the intervention group (r = 0.36 vs. 0.55), and some individual observations were discordant (**Supplementary Data 4**). For example, one control participant reported a 17.44% reduction in waist circumference despite a 1.02% increase in body weight at week 12. Such observations merit verification against the original records but do not, by themselves, demonstrate measurement error. Waist measurements may be particularly sensitive to tape placement, tension, posture, and breathing. Measurement variability and the different aspects of body composition captured by weight and waist circumference may partly explain their differing patterns (**Figure 6C and D**).

The interim findings provide preliminary observations for further investigation, while the completed analysis did not establish a statistically significant between-group effect. Small interim samples, baseline imbalances, variation in participant composition and follow-up availability, and possible behavioral differences offer plausible explanations for the discrepancy, but their individual contributions remain unresolved. Future studies incorporating balanced groups, standardized measurements, systematic adherence assessment, and blinded, placebo-controlled comparisons would help determine whether the numerical patterns reflect reproducible effects. Longer follow-up would also be needed to evaluate sustained responses and long-term safety.

Future studies should use an adequately powered, randomized, double-blind, placebo-controlled design with equivalent dietary guidance in both groups, prespecified analyses, standardized outcome measurements, and systematic safety monitoring. Important baseline characteristics should be balanced or addressed through prespecified statistical adjustment, while diet, physical activity, and adherence should be monitored throughout follow-up. Longer studies involving broader BMI ranges, including obesity, and participants with impaired glucose metabolism would help assess the durability and generalizability of any benefits. Dose optimization should be informed by validated pharmacokinetic and pharmacodynamic measurements to determine whether orally administered pro-DALAP produces sufficient systemic exposure to intact, active DALAP.

Overall, the experimental findings support yeast-based expression of pro-DALAP, preservation of host growth under the conditions evaluated, and functional receptor activation following peptide processing. The clinical study provides preliminary information on participant experience and short-term tolerability. Modest reductions in body weight and an overall downward trend were observed in the intervention group, although the between-group difference at week 12 was not statistically significant. Further investigation is needed to determine whether the platform’s demonstrated biological activity translates into reproducible clinical benefits following oral administration.

## Conclusion

The current study suggests that EV1-Peptide, a formulation containing yeast expressing pro-DALAP and serving as the active ingredient in the commercial product Evolv GLP-1, was generally well tolerated over the 12-week study period. Although between-group differences in the primary endpoints did not reach statistical significance, the observed trends, favorable safety profile within the study period, and positive participant feedback support further clinical evaluation using an optimized study design.

## Data Availability

Data supporting the findings of this study are included in the manuscript and supplementary materials. Additional data are available from the corresponding author upon reasonable request, subject to participant confidentiality and applicable consent restrictions.

## Acknowledgments

We thank Rebecca McCarthy, CEO and co-founder of BioDrive, Inc., and Olivia Ravlin, Chief Marketing Officer of BioDrive, Inc., for their assistance in coordinating the human clinical study. We also thank Citruslabs for conducting the randomized controlled trial, ITR Laboratories Canada Inc. for conducting the intravenous tolerability and maximum tolerated dose study in rats, and Eurofins DiscoverX for conducting the receptor activation assays.

## Declarations

### Author Contributions

Cory Henderson and Kwang-Chul Kwon conceptualized the study and coordinated the animal study conducted by ITR Laboratories Canada Inc. and the receptor activation assays conducted by Eurofins DiscoverX. Kwang-Chul Kwon established the Western blotting and densitometric quantification methods, performed Western blotting, densitometric analyses, furin cleavage assays, and GM1-binding assays, analyzed the human clinical trial data, organized and curated the study data, prepared the figures, and wrote the original manuscript. Cory Henderson reviewed and edited the manuscript. Olga Gaidarenko trained and supervised Nathan Chang in yeast engineering and experimental analyses. Together, Olga Gaidarenko and Nathan Chang generated all engineered yeast strains, including yeast transformation, genomic DNA extraction, and PCR screening to verify targeted genomic modifications. They also performed Western blotting, densitometric analyses, and yeast growth measurements. Olga Gaidarenko additionally performed microscopic examination of the yeast strains. All authors reviewed and approved the final manuscript.

### Data Availability

Data supporting the findings of this study are provided in the manuscript and supplementary materials. Additional de-identified individual participant data and the clinical study protocol may be made available by the corresponding author upon reasonable request, subject to applicable participant consent, confidentiality requirements, and sponsor approval.

### Ethics Approval

The human clinical trial conducted by Citruslabs was approved by Argus Independent Review Board (AIRB; Tucson, Arizona) on December 6, 2024 (protocol No. 20692). All participants provided written informed consent before undergoing any study-specific procedures.

### Consent for Publication

Not applicable. The manuscript presents aggregate results and de-identified individual participant data and contains no information identifying individual participants.

