## Supplementary Figure 1 for "A Yeast-Based Platform for Oral Delivery of a Dual GLP-1/GIP Receptor Agonist: Engineering and Preliminary Clinical Evaluation"

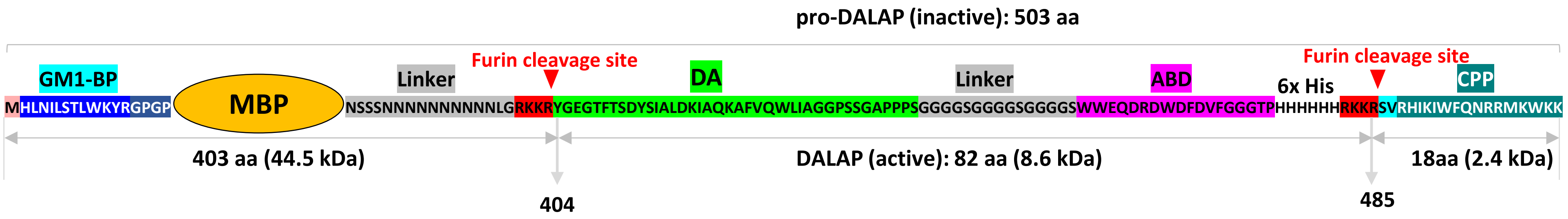


[1]MHLNILSTLWKYRGPGPKIEEGKLVIWINGDKGYNGLAEVGKKFEKDT
GIKVTVEHPDKLEEKFPQVAATGDGPDIIFWAHDRFGGYAQSGLLAEIT
PDKAFQDKLYPFTWDAVRYNGKLIAYPIAVEALSLIYNKDLLPNPPKTW
EEIPALDKELKAKGKSALMFNLQEPYFTWPLIAADGGYAFKYENGKYDI
KDVGVDNAGAKAGLTFLVDLIKNKHMNADTDYSIAEAAFNKGETAMT
INGPWAWSNIDTSKVNYGVTVLPTFKGQPSKPFVGVLSAGINAASPNK
ELAKEFLENYLLTDEGLEAVNKDKPLGAVALKSYEEELVKDPRIAATMEN
AQKGEIMPNIPQMSAFWYAVRTAVINAASGRQTVDEALKDAQTNSSS
NNNNNNNNNNLGRKKR[404]YGEGTFTSDYSIALDKIAQKAFVQWLIAGGP
SSGAPPPSGGGGSGGGGSGGGGSWWEQDRDWDFDVFGGGTPHH
HHHHRKK[485]RSVRHIKIWFQNRRMKWK[503]K

**Supplementary Figure 1. Domain organization and complete amino acid sequence of pro-DALAP.** The upper schematic illustrates the modular organization of pro-DALAP (503 amino acids), comprising an N-terminal GM1-binding peptide (GM1-BP), maltose-binding protein (MBP), linker sequences, the dual GLP-1/GIP receptor agonist peptide (DA), an albumin-binding domain (ABD), a hexahistidine tag (6×His), and a C-terminal cell-penetrating peptide (CPP). Two furin cleavage sites are highlighted in red, with arrowheads indicating the proposed cleavage positions. Cleavage is designed to release active DALAP (82 amino acids; approximately 8.6 kDa) from the inactive precursor, separating it from the N-terminal fragment (403 amino acids; approximately 44.5 kDa) and C-terminal fragment (18 amino acids; approximately 2.4 kDa). The complete amino acid sequence is shown below, with colors corresponding to the domains in the schematic. Selected residue positions are numbered.
