## Supplementary Figure 2 for "A Yeast-Based Platform for Oral Delivery of a Dual GLP-1/GIP Receptor Agonist: Engineering and Preliminary Clinical Evaluation"

**A**

| Blood Biomarker | Week 6 | | | | | Week 12 | | | | |
|---|---|---|---|---|---|---|---|---|---|---|
| | Intervention (n=31) | | Control (n=24) | | p-value | Intervention (n=33) | | Control (n=27) | | p-value |
| | Average | STD | Average | STD | | Average | STD | Average | STD | |
| Hemoglobin A1c (%) | **0.26** | 3.79 | **-0.62** | 4.3 | 0.999 | **0.73** | 3.77 | **-1.96** | 5.22 | 0.0486 |

**B**

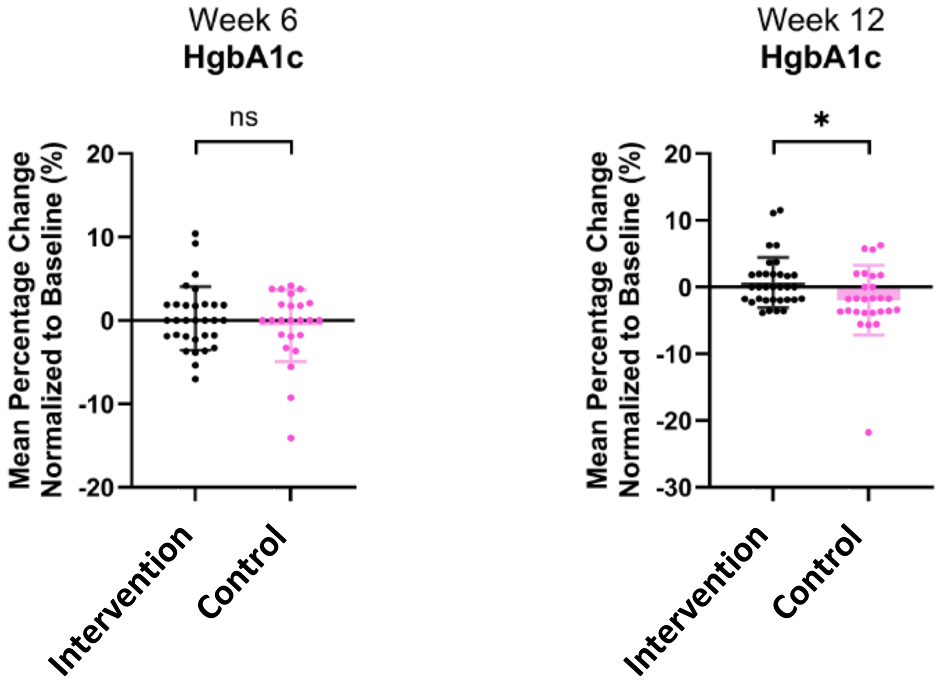


**Supplementary Figure 2. Percentage changes in hemoglobin A1c (HbA1c) at weeks 6 and 12. (A)** Mean percentage changes from baseline, standard deviations (SD), and between-group p-values for the test and control groups. **(B)** Corresponding dot plots showing individual participant values, with group means and SD. Negative values indicate reductions from baseline. Sample sizes were 31 and 24 for the test and control groups, respectively, at week 6, and 33 and 27 at week 12. Test, pro-DALAP supplementation group; Control, dietary advice group. ns, not significant ($p \geq 0.05$); $*p < 0.05$.
