## Supplementary Figure 3 for "A Yeast-Based Platform for Oral Delivery of a Dual GLP-1/GIP Receptor Agonist: Engineering and Preliminary Clinical Evaluation"

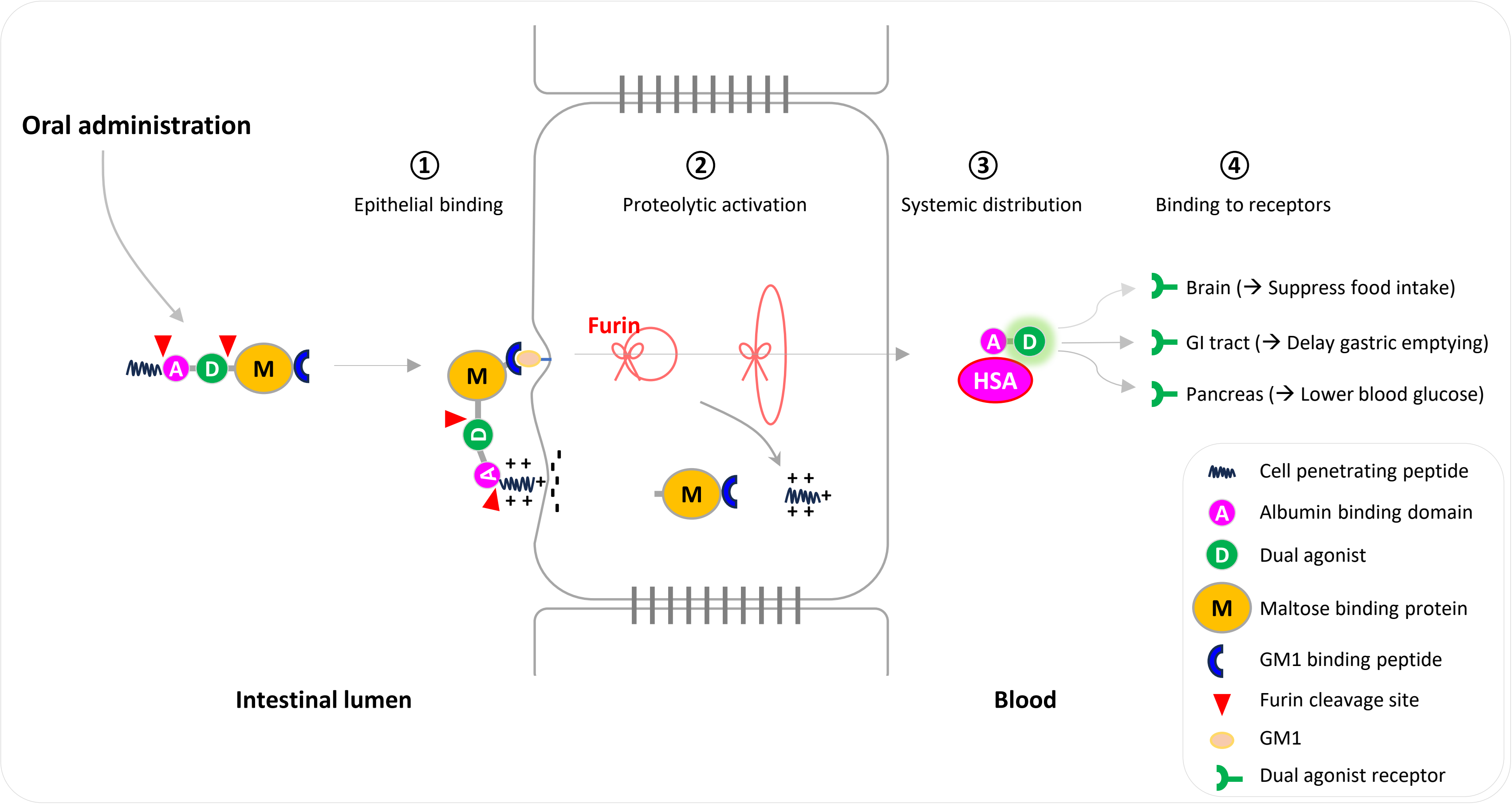


**Supplementary Figure 3. Proposed mechanism of oral pro-DALAP delivery and action.** (1) Following release from the enteric-coated formulation in the intestinal lumen, the terminal targeting peptides of pro-DALAP are proposed to promote epithelial membrane binding and uptake. (2) Furin-mediated cleavage during transcytosis is proposed to release active DALAP. (3) Binding of DALAP to human serum albumin through its albumin-binding domain is expected to prolong its circulating half-life. (4) Following systemic distribution, DALAP is proposed to activate GLP-1 and GIP receptors, potentially influencing appetite, gastric emptying, and glucose regulation.
